# Lung transcriptomics of radiologic emphysema reveal barrier function impairment and macrophage M1-M2 imbalance

**DOI:** 10.1101/2022.10.21.22281369

**Authors:** Robin Lu, Andrew Gregory, Rahul Suryadevara, Zhonghui Xu, Dhawal Jain, Brian D. Hobbs, Noah Lichtblau, Robert Chase, Edwin K. Silverman, Craig P. Hersh, Peter J. Castaldi, Adel Boueiz, the COPDGene investigators

## Abstract

**Rationale:** While many studies have examined gene expression in lung tissue, the gene regulatory processes underlying emphysema are still not well understood. Finding efficient non-imaging screening methods and disease-modifying therapies has been challenging, but knowledge of the transcriptomic features of emphysema may help in this effort.

**Objectives:** Our goals were to identify emphysema-associated biological pathways through transcriptomic analysis of bulk lung tissue, to determine the lung cell types in which these emphysema-associated pathways are altered, and to detect unique and overlapping transcriptomic signatures in blood and lung.

**Methods:** Using RNA-sequencing data from 456 samples in the Lung Tissue Research Consortium and 2,370 blood samples from the COPDGene study, we examined the transcriptomic features of computed tomography quantified emphysema. We also queried lung single-cell RNA-sequencing data to identify cell types showing COPD-associated differential expression of the emphysema pathways found in the bulk analyses.

**Measurements and Main Results:** In the lung, 1,055 differentially expressed genes and 29 dysregulated pathways were significantly associated with emphysema. We observed alternative splicing of genes regulating NF-κB and cell adhesion and increased activity in the TGF-β and FoxO signaling pathways. Multiple lung cell types displayed dysregulation of epithelial barrier function pathways, and an imbalance between pro-inflammatory M1 and anti-inflammatory M2 macrophages was detected. Lung tissue and blood samples shared 251 differentially expressed genes and two pathways (oxidative phosphorylation and ribosomal function).

**Conclusions:** This study identified emphysema-related changes in gene expression and alternative splicing, cell-type specific dysregulated pathways, and instances of shared pathway dysregulation between blood and lung.

**AT A GLANCE COMMENTARY:** *Scientific Knowledge on the Subject:* Prior studies have investigated the transcriptomic characteristics of emphysema and its associated biological pathways. However, less is known about alternative splicing mechanisms and cell-type specific transcriptional patterns in emphysema. Additionally, a comparison between dysregulated genes and pathways in blood and lung tissues is needed to better understand the utility of non-invasive diagnostic and prognostic tools for emphysema.

*What This Study Adds to the Field:* Using lung samples from the Lung Tissue Research Consortium (LTRC) and blood samples from the COPDGene study, we performed differential gene and alternative splicing association analyses for CT-quantified emphysema. We then queried a previously published lung tissue single-cell RNA-sequencing atlas of COPD patients and controls to determine lung cell-type specific expression patterns of the biological pathways identified from the bulk analyses. We demonstrated that multiple pathways, including oxidative phosphorylation and ribosomal function processes, were enriched in both blood and lung tissues. We also observed that in COPD, oxidative phosphorylation was downregulated in pro-inflammatory (M1) macrophages and upregulated in anti-inflammatory (M2) macrophages. Additionally, other immunity-related cell types, including plasma cells, natural killer cells, and T lymphocytes, were linked to epithelial barrier function, such as the Rap1, adherens junction, and TGF-β signaling pathways.

## INTRODUCTION

Chronic obstructive pulmonary disease (COPD) is a major source of morbidity and mortality (1). Emphysema, an important COPD phenotype, has been shown to be independently associated with elevated risk for cardiovascular disease, lung cancer, and mortality (2-4). Finding efficient non-imaging screening methods and targeted therapies may be aided by knowledge of the transcriptomic characteristics of emphysema. Although emphysema has been linked to genes involved in transforming growth factor beta (TGF-β) signaling (5, 6), B-cell mediated immunity (5, 7, 8), and hypoxia (8-10), our understanding of emphysema-associated alternative splicing mechanisms and cell-type specific biological pathways from human lung tissue is still limited.

Earlier transcriptomic studies of emphysema were limited in their sample sizes or the scope of the panel of genes evaluated and have not investigated alternative splicing extensively. In addition, although oxidative stress (11, 12) and cellular senescence (13, 14) have been associated with emphysema, it is unclear whether other biological processes may also be at play. It is also less obvious which cell types exhibit pathway dysregulation in the lungs of subjects with emphysema compared to controls, although it is widely recognized that cell types such as neutrophils (15, 16) and T-lymphocytes (17, 18) are drivers of the disease. Comparing dysregulated genes and pathways in blood and lung tissues is also necessary to better understand the utility of non-invasive diagnostic and prognostic tools for emphysema.

In the present study, we used genome-wide RNA-sequencing (RNA-seq) data from lung tissue samples from the Lung Tissue Research Consortium (LTRC) to identify the genes and alternative splicing mechanisms that are associated with computed tomography (CT)-quantified emphysema. We then queried a previously published single-cell RNA-seq atlas of lung tissues of COPD patients and controls to determine which cell types show significant associations to the identified emphysema pathways. We lastly compared emphysema-associated transcriptomic associations in LTRC lung tissue samples to emphysema-associations from whole blood samples in the COPD Genetic Epidemiology (COPDGene) study. We hypothesized that there would be significant emphysema-associated transcriptomic biomarkers and pathways identified from lung tissue, cell-type specific signatures, and important similarities and differences between lung tissue and whole blood.

## METHODS

### Study description

We obtained lung tissue samples from the NHLBI Lung Tissue Research Consortium (LTRC) (https://www.nhlbi.nih.gov/science/lung-tissue-research-consortium-ltrc, https://biolincc.nhlbi.nih.gov/studies/ltrc/). Details regarding subject recruitment has been previously published (19). Subjects included smokers and non-smokers over the age of 21 who had undergone surgical lung biopsy, lung volume reduction surgery, lung transplantation, or lung nodule/mass resection. These subjects cover the entire spectrum of the Global Initiative for Chronic Obstructive Lung Disease (GOLD) spirometric grading system (20). We excluded subjects missing the following clinical data: CT-quantified emphysema, CT scanner model, forced expiratory volume in one second (FEV_1_), body mass index (BMI), current smoking status, and pack-years of smoking. We also excluded subjects with idiopathic pulmonary fibrosis (IPF). IPF was defined according to the American Thoracic Society/European Respiratory Society guidelines as a consensus clinical diagnosis of IPF or a pathologic diagnosis of usual interstitial pneumonia or honeycomb lung in the absence of a clinical diagnosis of another interstitial lung disease (21).

All analyses conducted on LTRC were also completed in previously published analyses using whole blood RNA-seq data from the COPDGene study. The COPDGene study is a longitudinal study investigating the epidemiologic and genomic characteristics of COPD. This study included 10,371 smokers from 21 U.S. clinical institutions centers, representing the full spectrum of lung health, who were between the ages of 45 and 80 and had at least ten pack-years of lifetime cigarette smoking history (NCT00608764, www.copdgene.org) (22). COPDGene continues to collect longitudinal data on study participants at five-year intervals. Each study visit collected spirometry data, questionnaires, and chest CT scans using a standard protocol. Whole blood RNA-seq was obtained at Visit 2.

All subjects provided informed consent, and all clinical sites received institutional review board approval.

### Emphysema quantification

The Analyze 8.1 (www.analyzedirect.com) (23) and Thirona (www.thirona.eu) softwares quantified emphysema in LTRC and COPDGene, respectively. Emphysema was measured as the Hounsfield units (HU) at the 15^th^ percentile of the CT density histogram at end-inspiration (Perc15 density) (24, 25). In COPDGene, the Perc15 density values were corrected for the inspiratory depth variations (adjusted Perc15 density). Both Perc15 and adjusted Perc15 are given as the HU + 1000. The lower the Perc15 or adjusted Perc15 values are, i.e., the closer to −1,000 HU, the more CT-quantified emphysema is present.

### Differential expression and usage analyses

We used the limma-voom linear modeling approach (as implemented in limma v3.46.0) to test for the associations between emphysema and whole blood RNA transcripts (26, 27). While differential expression refers to the change in the *absolute* expression levels of a feature, differential usage captures alternative splicing and refers to the change in the *relative* expression levels of the isoforms/exons within a given gene. The concepts of differential expression and usage with a discrete variable are naturally extended to a continuous variable where the changes in expression and usage mean changes in the absolute and relative rate of association with emphysema. All models were adjusted for age, race, sex, pack-years of smoking, current smoking status, FEV_1_, CT scanner model, and library preparation batch. The LTRC analysis was also adjusted for BMI. The COPDGene analysis was also adjusted for CBC cell count proportions.

### Pathway analysis

We used the egsea software (v1.18.1) to perform gene set enrichment analysis (GSEA) on the gene sets obtained from the differential gene expression (DGE) analysis (28). The KEGG pathway gene sets were utilized as the reference for annotated gene sets, and the CAMERA base approach was employed (29, 30). CAMERA is a competitive gene set test approach that employs estimated inter-gene correlation to adjust the gene set test statistic (30). The directionality of a pathway was ascertained by counting the number of upregulated and downregulated genes in the gene set and taking the direction of the majority (28). The genes corresponding to these pathways were mapped on the pathway maps given by the KEGG database using the pathview and gage libraries in R. Twenty-nine pathways in total were found to be dysregulated in our data, but five (Parkinson’s disease, Huntington’s disease, dorso-ventral axis formation, Alzheimer’s disease, and proteasome) were not mapped to the KEGG database. For easier visualization and interpretation, the reported gene log fold changes in the pathway diagrams were multiplied by −100 so that positive log fold changes represent upregulated genes and negative log fold changes represent downregulated genes. Thus, the fold change estimates correspond to expression change per 100 HU decrease in lung density (increasing emphysema).

### Cell-type specificity analysis

We re-analyzed data from a previously published lung tissue single-cell RNA-seq experiment to identify enriched pathways from the bulk lung DGE analyses at cell type resolution (31). The dataset consists of 312,928 single cells from 78 human samples (28 control, 18 end-stage COPD, 32 IPF). The control samples in this cohort represented allograft rejected donors, and only control and end-stage COPD samples were used for this analysis, while keeping the author-provided cell type annotations.

A composite gene expression signature for the pathway members was summarized using the AddModuleScore function from the Seurat package (32). Briefly, the function calculates a normalized average expression for a set of genes belonging to a given pathway subtracted by an aggregate expression of control genes. All genes in the dataset were binned based on their expression levels. For each gene in the pathway, control genes were randomly selected from bins with matching expressions. This ensured similar distribution of the gene expression values in the control set. Finally, the activity of the pathway was defined by subtracting the averaged expression of control genes from the average expression of the member genes for each cell. We employed this background-corrected gene expression signature approach to calculate pathway activity scores for the 24 KEGG pathways identified using GSEA across every single cell in the single-cell dataset. We compared the pathway activity scores between controls and end-stage COPD for each cell type separately using the two-sample t-tests (33).

Next, we scored the specificity of a pathway across cell types using the pathway activity scores. The pathway activity scores showed a normal unimodal distribution. We, therefore, used an arbitrary cutoff of the mean plus two times the standard deviation to assess the activity of a given pathway in all cells. This stringent positive cutoff value allows us to look at cell types that strongly enrich a given pathway activity. The odds ratio was computed for the cells showing the pathway activity versus not showing activity in a given cell type. The P-values were calculated using the Fisher’s Exact Test (34). The negative log10 of the corrected P-values were capped at 100 and plotted in a heatmap using the ComplexHeatmap package (35).

### Statistical analyses

Data were reported as mean with standard deviations or counts with percentages. Upregulated versus downregulated genomic features or KEGG pathways were provided with respect to their relationships with emphysema (i.e., they have opposite directions for their associations with Perc15 or adjusted Perc15 density). Because Perc15 and adjusted Perc15 decrease with more severe emphysema, negative log fold change values represent features upregulated with emphysema, and positive log fold change values represent features downregulated with emphysema. The Benjamini-Hochberg method corrected for multiple comparisons using a threshold of significance of a false discovery rate (FDR) of 10% (36).

## RESULTS

### Study subjects

The study flow diagram is depicted in Figure 1, and the missingness of the pertinent covariates is shown in Figure E1. A total of 456 LTRC samples coming from 446 participants were included in this analysis. Eight subjects had multiple lung tissue samples taken on the same date. Of the 1,589 LTRC samples for which RNA-seq data were available, 508 had comprehensive clinical information including quantitative CT emphysema. We further excluded 52 samples with IPF. Table 1 provides an overview of the demographics and clinical characteristics of the included subjects. The majority of these subjects were non-Hispanic whites with a mean age of 64, a mean BMI of 28.1, and a balanced sex representation. 8.1% were current smokers with an average of 36 pack-years of smoking. With the exception of more smoking and slightly less spirometric impairment in the included subjects, there were no significant differences found between the included and excluded subjects (Table E1). The baseline characteristics of the LTRC and the COPDGene individuals were also comparable (Table E2), as well as the distributions of their respective Perc15 and adjusted Perc15 values (Figure E2).

**Table 1.**
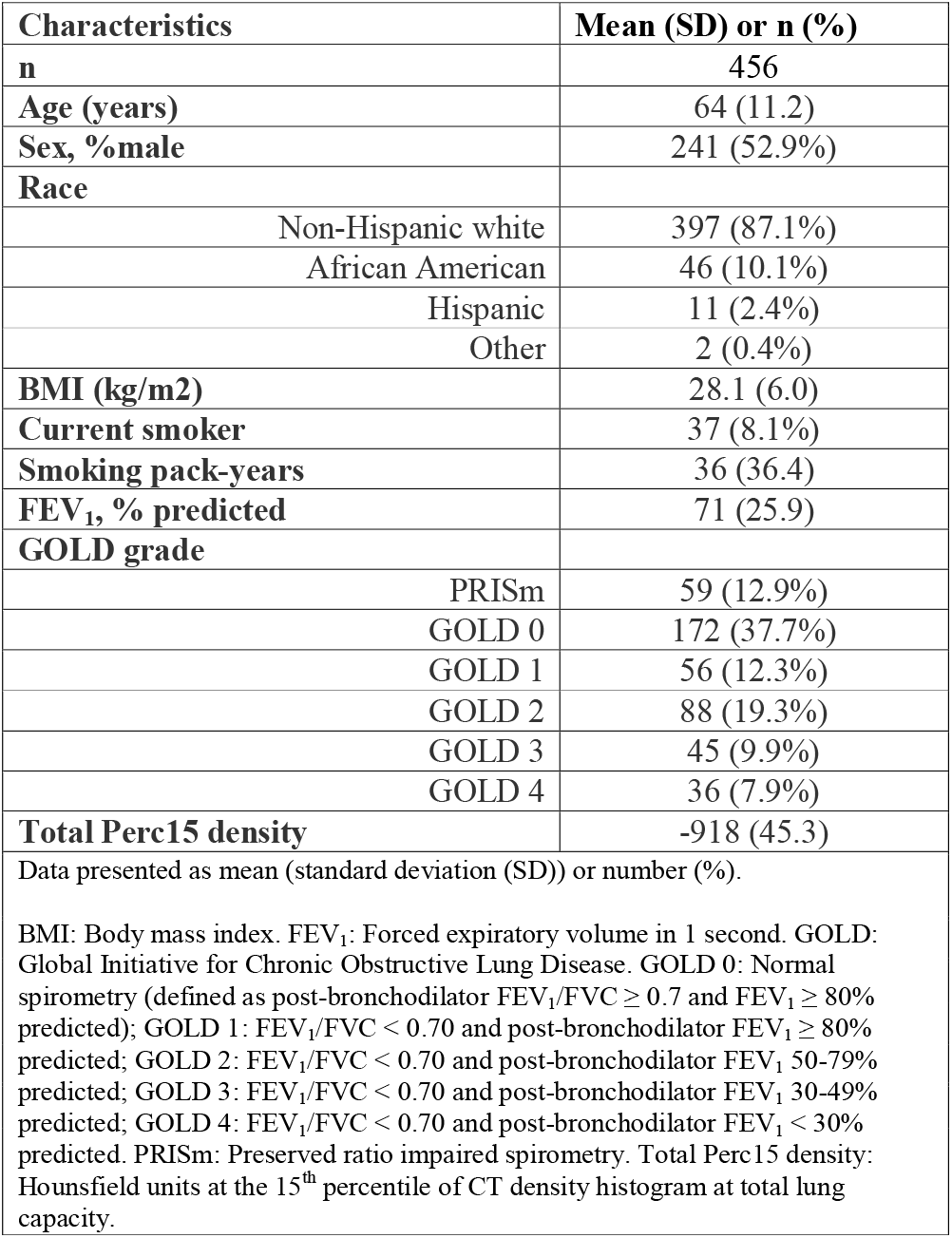
Baseline characteristics of the included Lung Tissue Research Consortium (LTRC) subjects.

**Figure 1.**
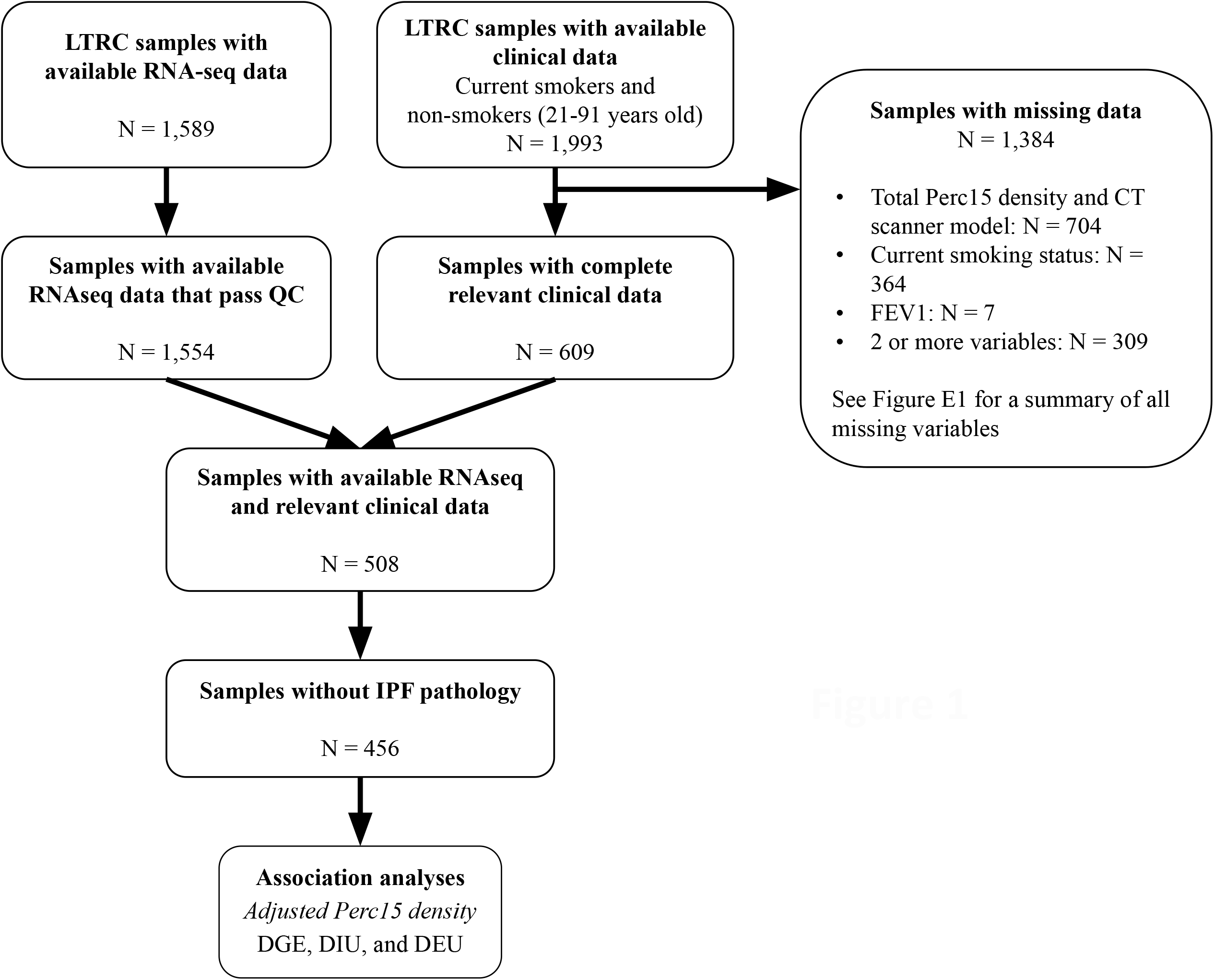
Study flow diagram. Abbreviations: BMI: Body mass index. FEV_1_: Forced expiratory volume in 1 second. IPF: Idiopathic pulmonary fibrosis. LTRC: Lung Tissue Research Consortium. QC: Quality control. RNA-seq: RNA sequencing. Total Perc15 density: Hounsfield units at the 15^th^ percentile of CT density histogram at total lung capacity.

### Differential gene expression in lung tissue

A total of 1,055 of the 17,353 genes evaluated in lung tissue samples achieved statistical significance at FDR 10% (Table E3). Of these significant genes, 632 were upregulated and 423 were downregulated with increasing emphysema. The top 20 most significant differentially expressed genes (DEGs) are listed in Table 2. Interestingly, the *FOXL1* and *F2RL2* genes, which had previously been found to be overexpressed in IPF lungs (37, 38), were downregulated in lung tissues with emphysema. Table E4 presents the 29 significantly dysregulated pathways found by performing gene set enrichment analysis on the identified DEGs. Table 3 reports ten noteworthy pathways that were selected for inclusion based on their biological relevance. There was enhanced activity in pluripotency pathways (TGF-β and forkhead box O (FoxO) signaling) and cell barrier function pathways (adherens junction, Rap1 signaling, and gap junction). Figure 2 displays KEGG pathway maps that show coordinated expression changes for TGF-β signaling, FoxO signaling, adherens junction, and Rap1 signaling, which have been previously found to be dysregulated in COPD or emphysema (39-44). Additional KEGG pathway maps are included in the supplemental materials for the remaining six significantly enriched pathways (Figure E3).

**Table 2.**
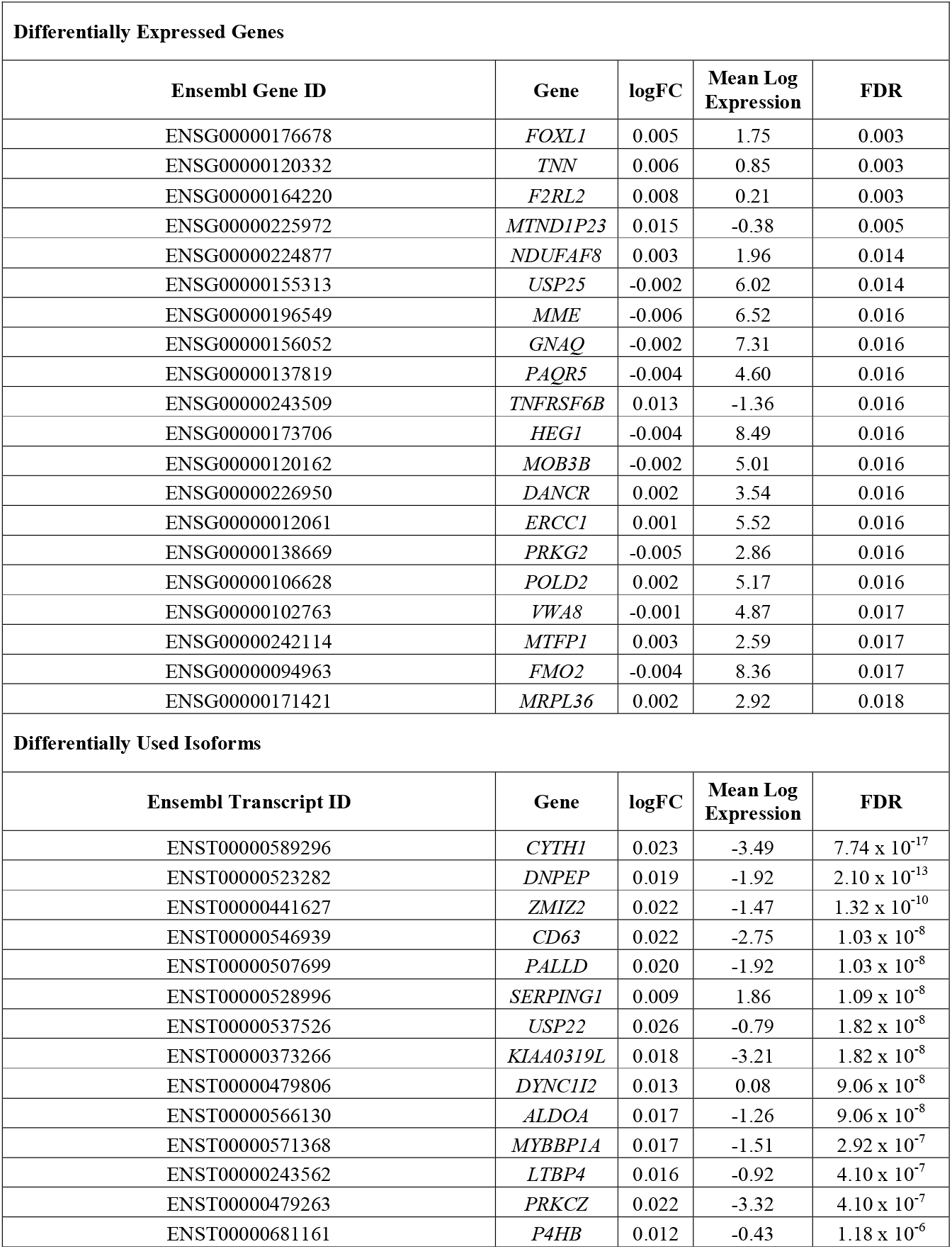
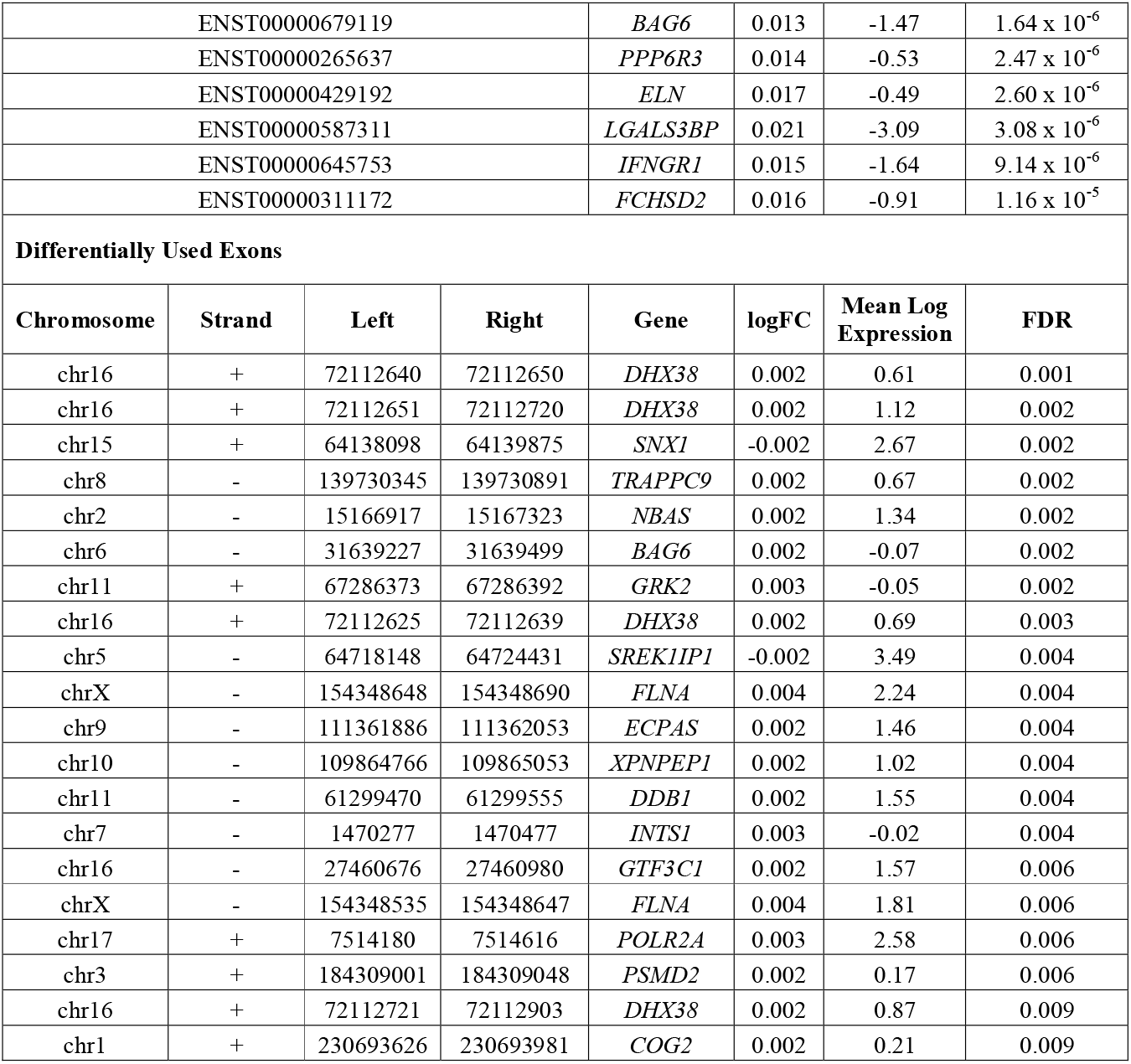
Top 20 differentially expressed or used features in lung tissue. logFC = log fold change (expression/usage change per Hounsfield unit); FDR = false discovery rate.

**Table 3.** Select significant pathways (FDR 10%) from the gene set enrichment analyses results using differentially expressed genes from lung tissue.

| Term Name | Number of genes annotated | Average logFC | FDR |
| --- | --- | --- | --- |
| Oxidative phosphorylation | 125 | 0.001 | $1.18 \times 10^{-4}$ |
| Signaling pathways regulating pluripotency of stem cells | 110 | -0.002 | $2.36 \times 10^{-2}$ |
| Adherens junction | 70 | -0.002 | $5.30 \times 10^{-2}$ |
| FoxO signaling pathway | 120 | -0.001 | $6.03 \times 10^{-2}$ |
| Regulation of actin cytoskeleton | 183 | -0.002 | $6.73 \times 10^{-2}$ |
| Jak-STAT signaling pathway | 109 | -0.002 | $7.57 \times 10^{-2}$ |
| Rap1 signaling pathway | 186 | -0.003 | $7.58 \times 10^{-2}$ |
| Fc epsilon RI signaling pathway | 62 | -0.001 | $7.58 \times 10^{-2}$ |
| TGF-beta signaling pathway | 76 | -0.002 | $8.61 \times 10^{-2}$ |
| Gap junction | 75 | -0.003 | $9.41 \times 10^{-2}$ |
| Upregulated versus downregulated pathways are provided with respect to their relationships with Perc15 density which is opposite to emphysema. A negative logFC value indicates a pathway upregulated with emphysema, and a positive logFC value indicates a pathway downregulated with emphysema. Pathways were reported based on relevance to lung disease. Average logFC = mean log fold change of genes in the pathway (gene expression change per Hounsfield unit); FDR = false discovery rate. |  |  |  |

**Figure 2.**
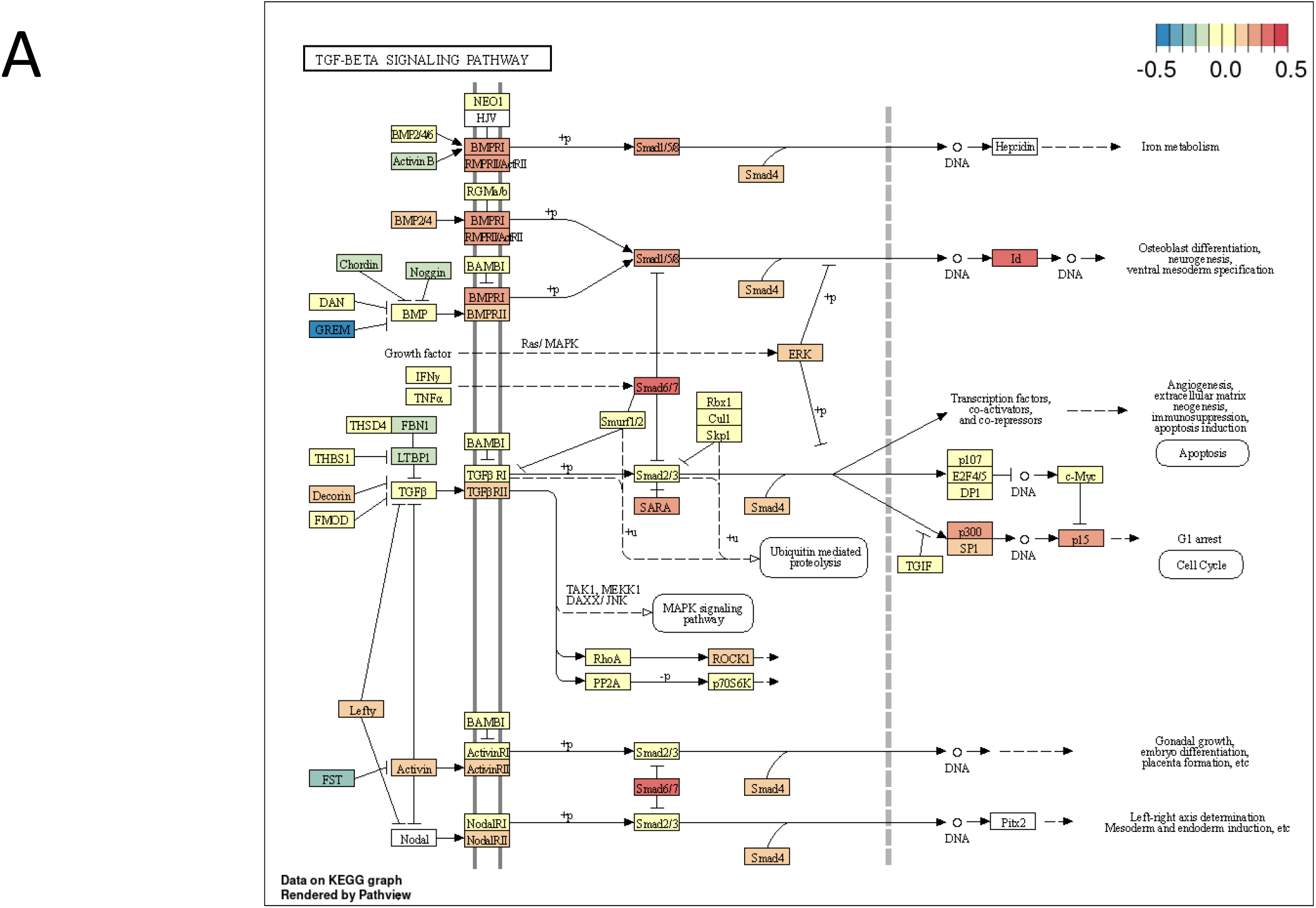
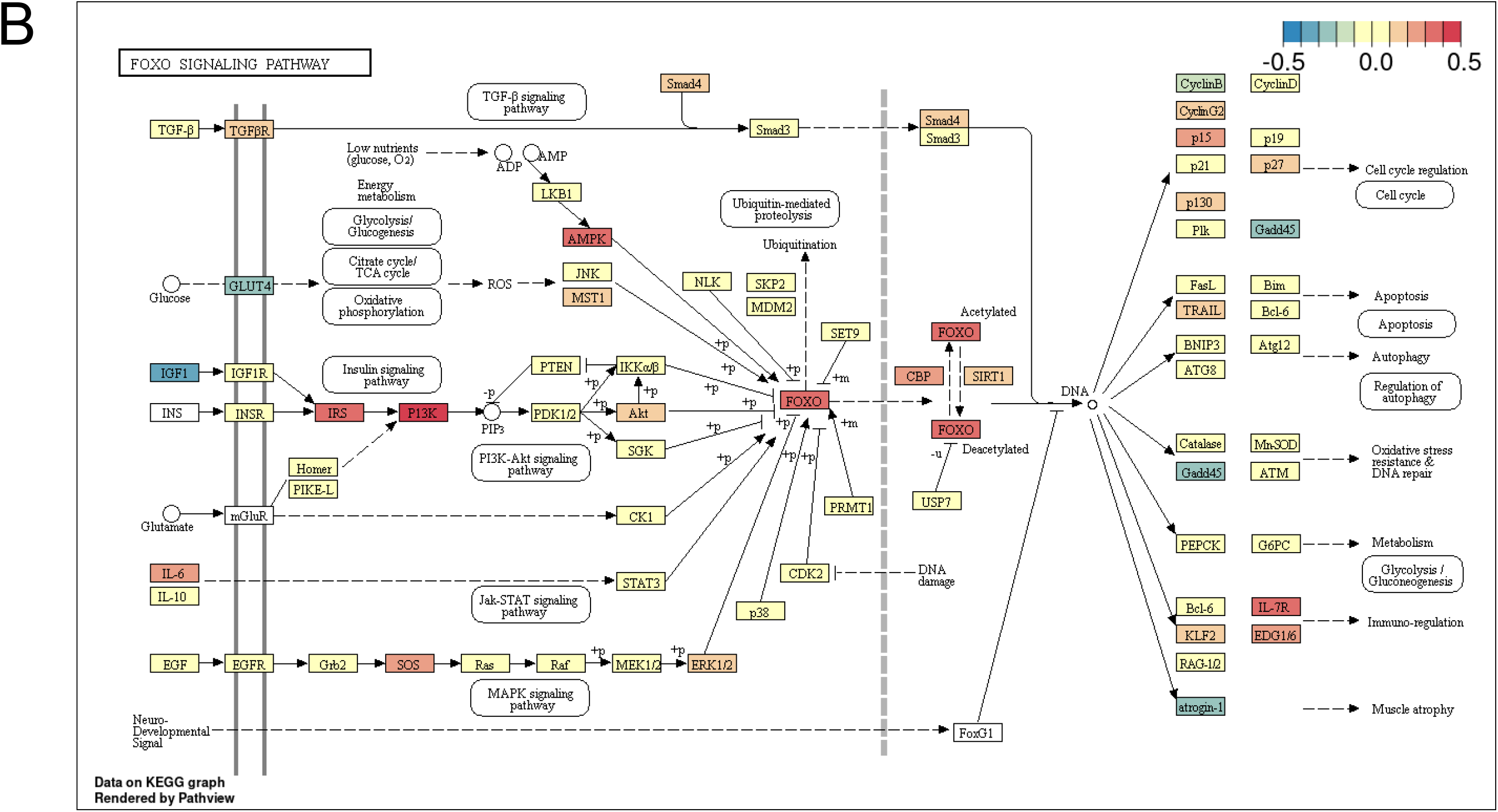
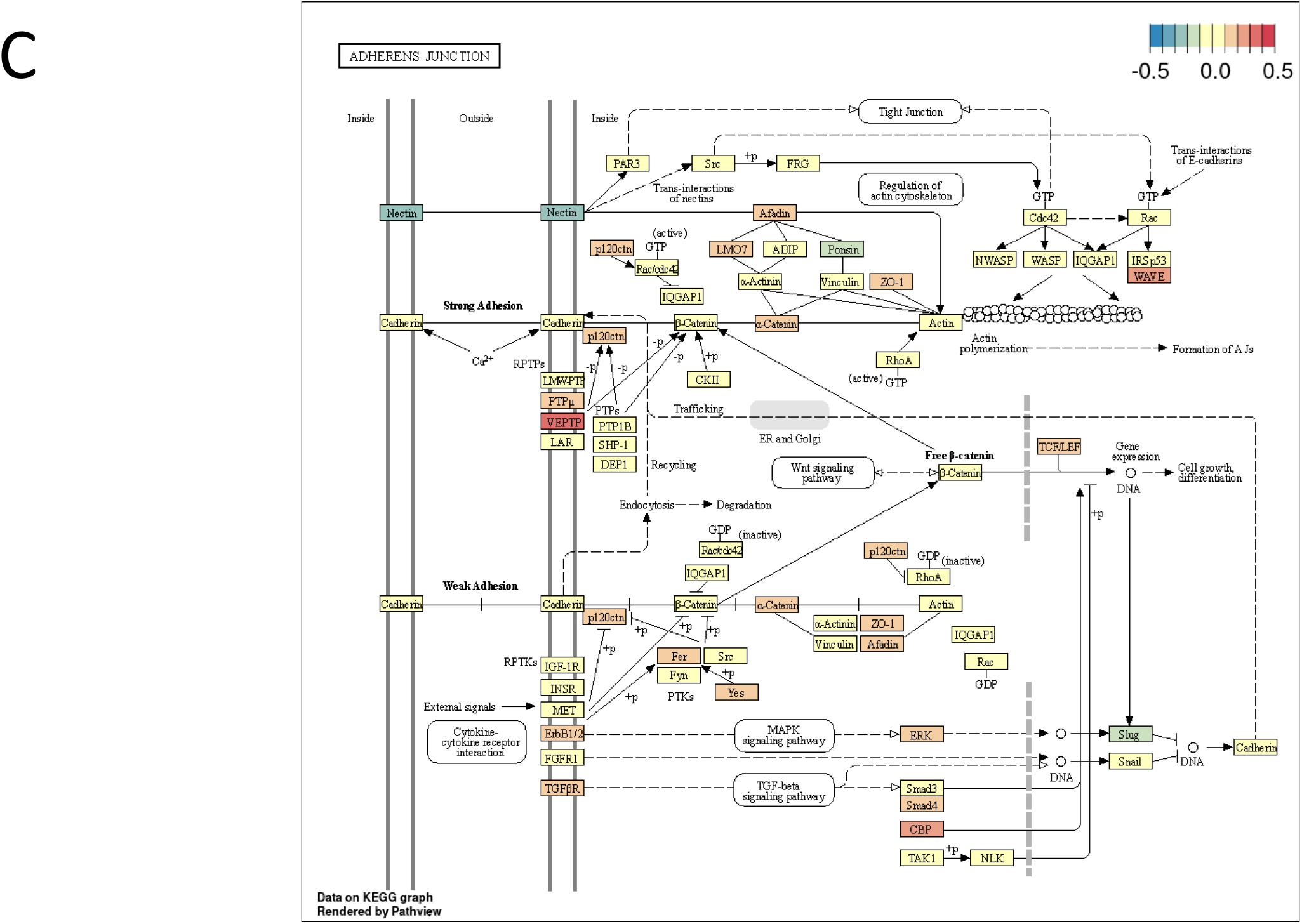
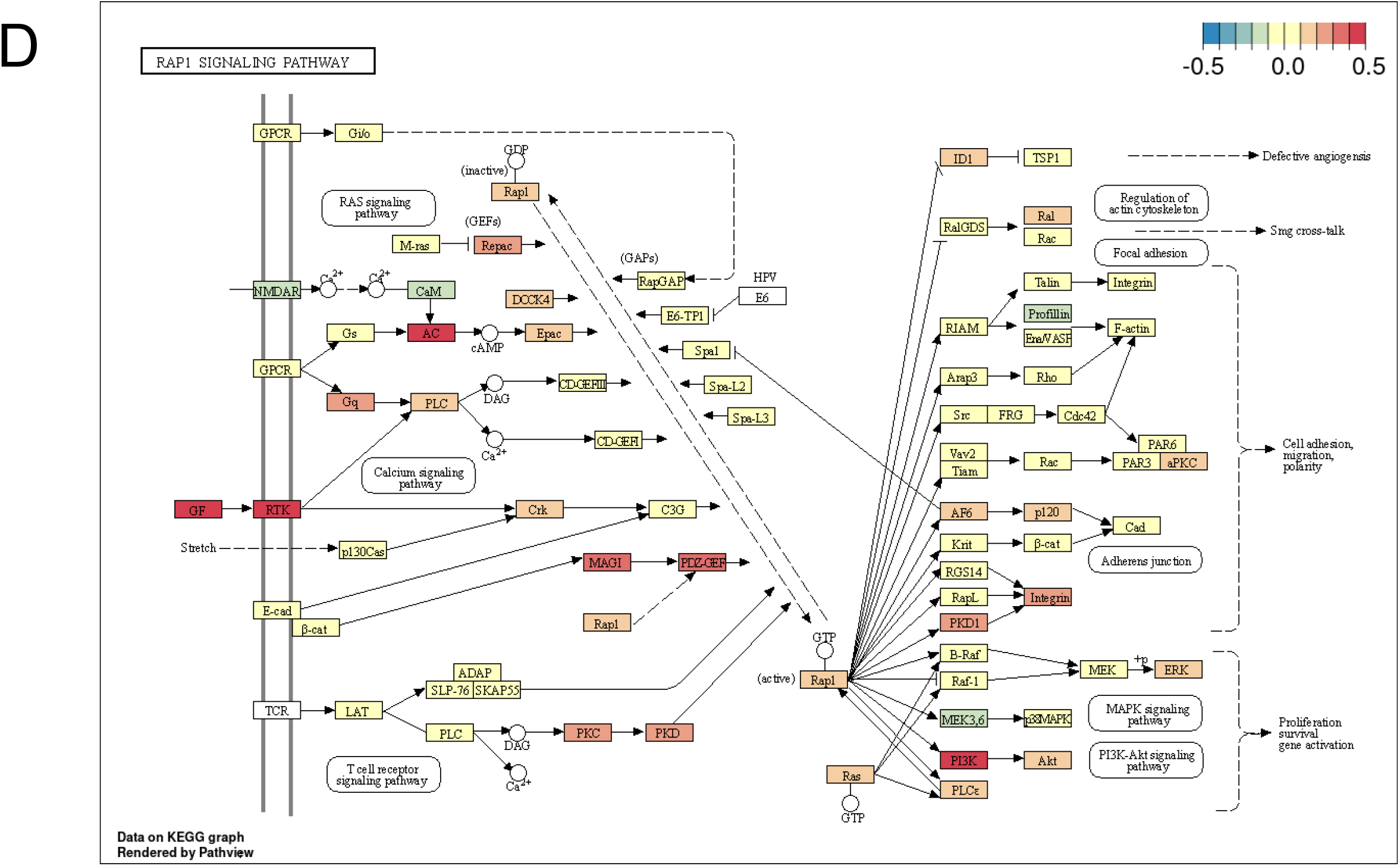
Kyoto Encyclopedia of Genes and Genomes (KEGG) pathway maps of the (**A**) TGF-β signaling pathway, (**B**) adherens junction, (**C**) Rap1 signaling pathway, and (**D**) FoxO signaling pathway reporting the effect size (log fold change) of all genes within each pathway in the 456 lung tissue samples from subjects in the Lung Tissue Research Consortium (LTRC). Emphysema was quantified by Hounsfield units at the 15^th^ percentile of chest CT density histogram at full inspiration (Perc15). The lower the Perc15 values are, i.e. the closer to −1,000 HU, the more CT-quantified emphysema is present. Gene log fold changes were multiplied by −100 so that positive log fold changes represented upregulated genes and negative log fold changes represented downregulated genes. Red represents upregulated genes and blue represents downregulated genes.

### Differential isoform and exon usage in lung tissue

Differential isoform usage (DIU) and differential exon usage (DEU) analyses were performed to understand emphysema-associated alternative splicing changes in lung tissue. A summary of the top twenty DUIs and DUEs can be found in Table 2.

Of the 41,891 isoforms evaluated, 730 isoforms were significantly associated with emphysema *(FDR 10%)* (Table E5). Of these significant isoforms, 271 were upregulated and 459 were downregulated. Mapping these isoforms to their corresponding genes revealed that 6.9% (41/598) of them were significant in the DGE analysis (Figure 3). These corresponding genes included MYB binding protein 1a (*MYBBP1A*) and protein kinase C zeta (*PRKCZ*), regulators of nuclear factor kappa-light-chain-enhancer of activated B cells (NF-κB) (45, 46). Additionally, there were significant isoforms mapping to CD63 and paladin (*PALLD*), which are involved in cell adhesion and cell-cell junctions (47, 48). An isoform of interferon gamma receptor 1 (*IFNGR1*) was also significantly associated with emphysema.

**Figure 3.**
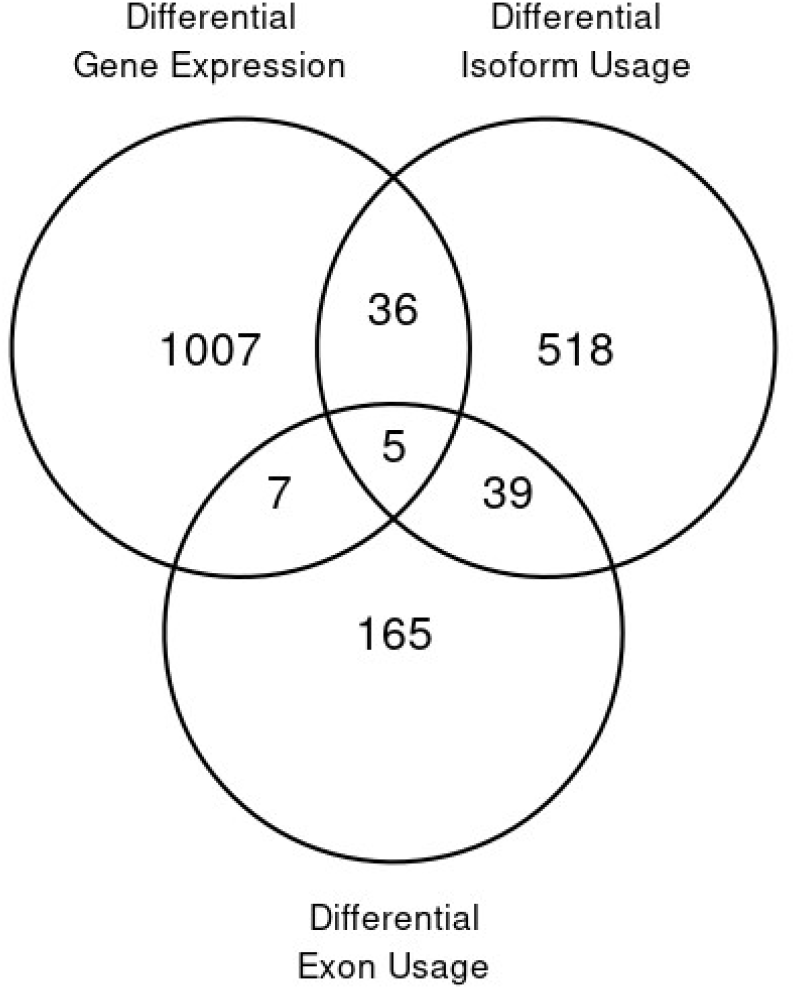
Venn diagram of the number of significant emphysema-associated genes from the differential gene expression, differential isoform usage, and differential exon usage models from lung tissue.

Of the 162,747 exons evaluated, 285 were significantly associated *(FDR 10%)* with emphysema with 24 being upregulated and 261 being downregulated (Table E6). After mapping these exons to their respective genes, we found that 5.6% (12/216) of the genes were differentially expressed (Figure 3). One of the top significantly associated exons corresponded to *TRAPPC9*, which codes for a protein that regulates NF-κB activation (49). We also identified exons within *SNX1, NBAS*, and *COG2*, which are genes involved in intracellular transport (50-52).

### Identification of cell-type specific pathways

To identify lung cell types showing the highest expression levels of emphysema associated pathways, we computed the odds ratio for each pathway and cell type using the pathway activity scores. The odds ratio was computed for the cells showing pathway activity versus not showing activity in a given cell type and for the remainder of the data. The heatmap revealed distinct clusters of the most strongly enriched pathways (Figure 4A). The first cluster included the phosphatidylinositol signaling, spliceosome, long term potentiation, and ribosome biogenesis pathways which were enriched in T cells and cytotoxic T cells. In the second cluster, the Fc-epsilon RI signaling pathway, platelet activation, Rap1 signaling pathway, and FoxO signaling pathways were enriched in both classical and non-classical monocytes. Adherens junction and TGF-β signaling pathway which were enriched in multiple cell types, including pulmonary cells, vascular endothelial cells, alveolar cells, and epithelial cells, made up the third major cluster.

**Figure 4.**
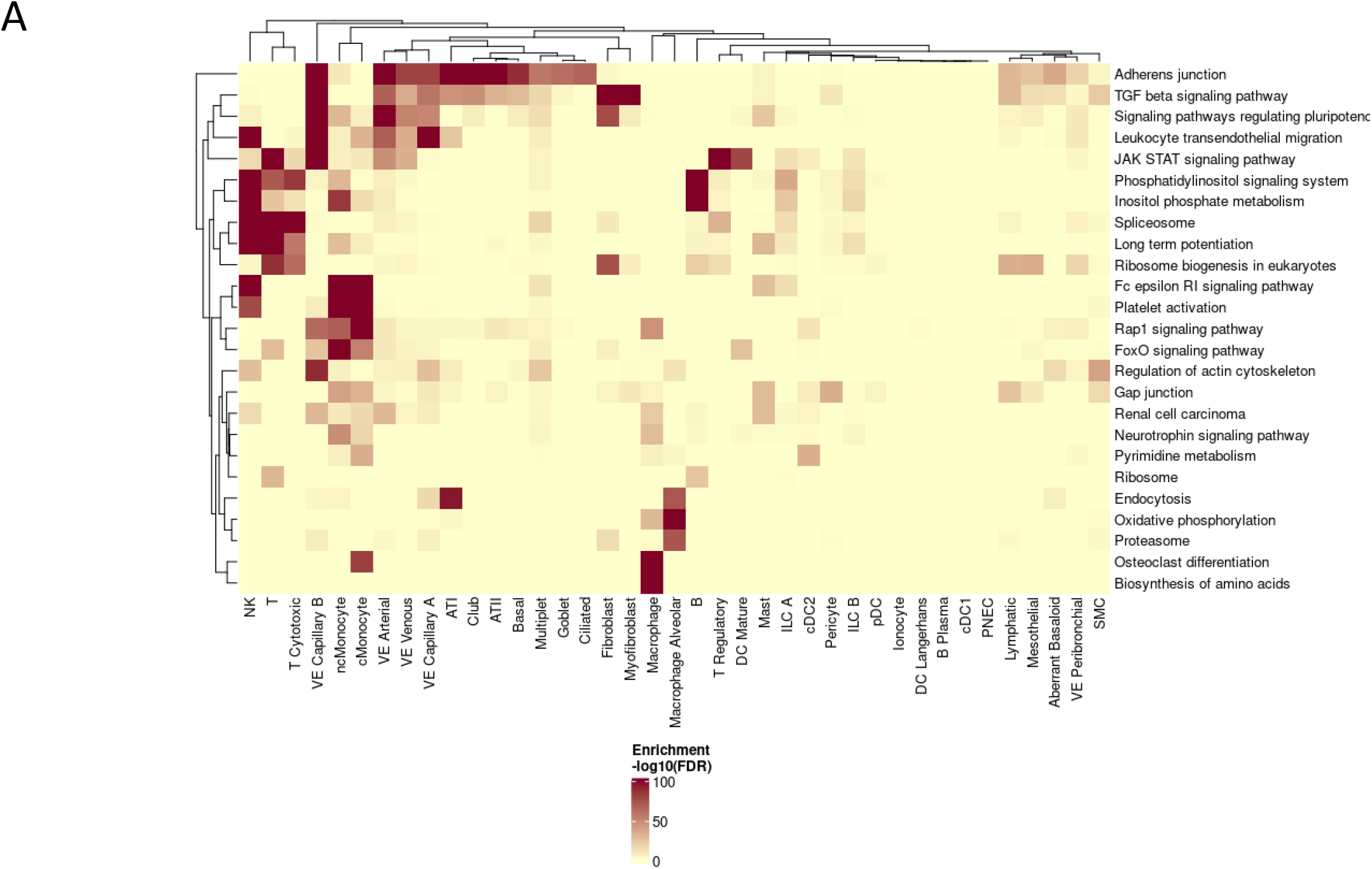
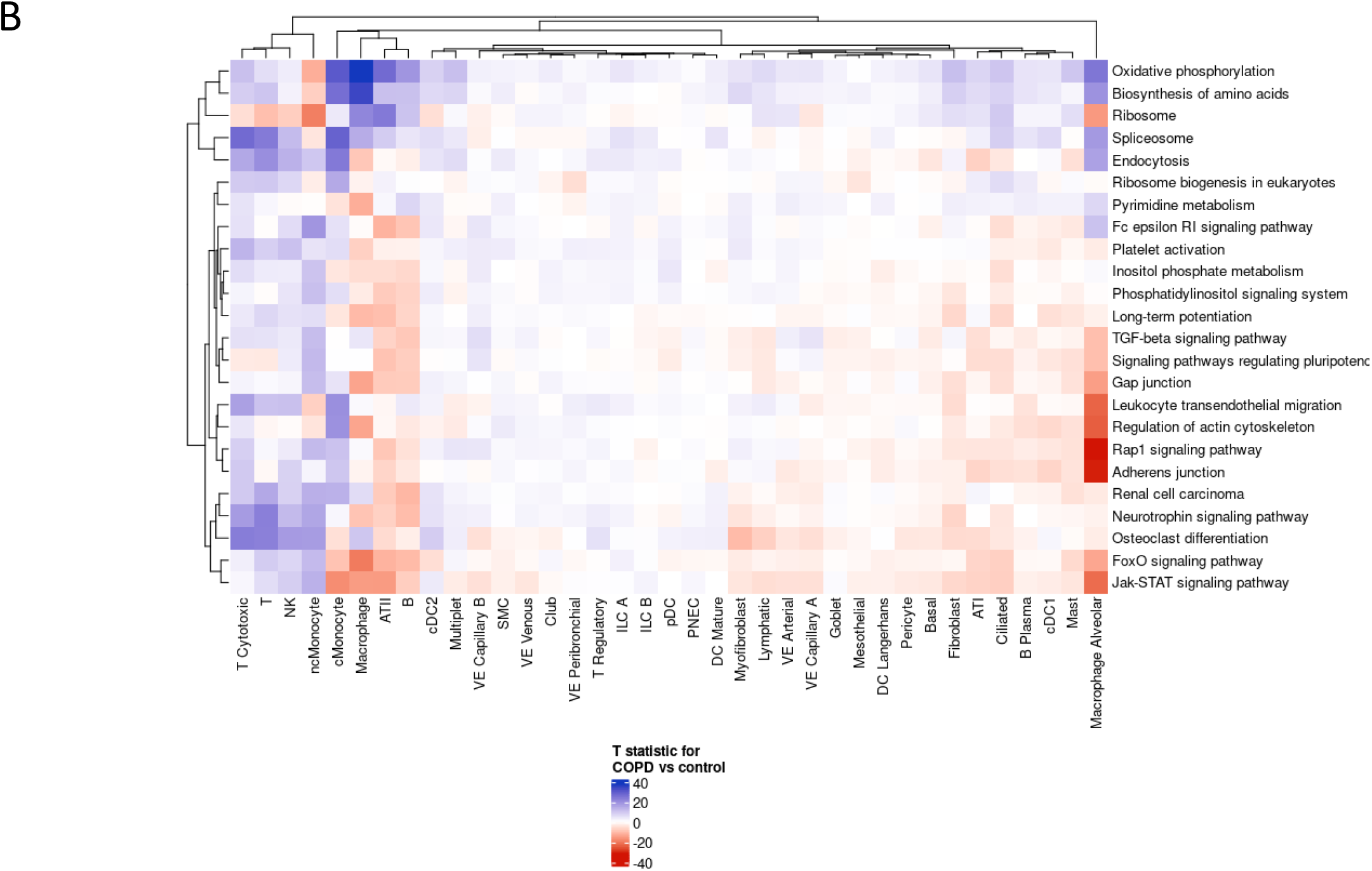
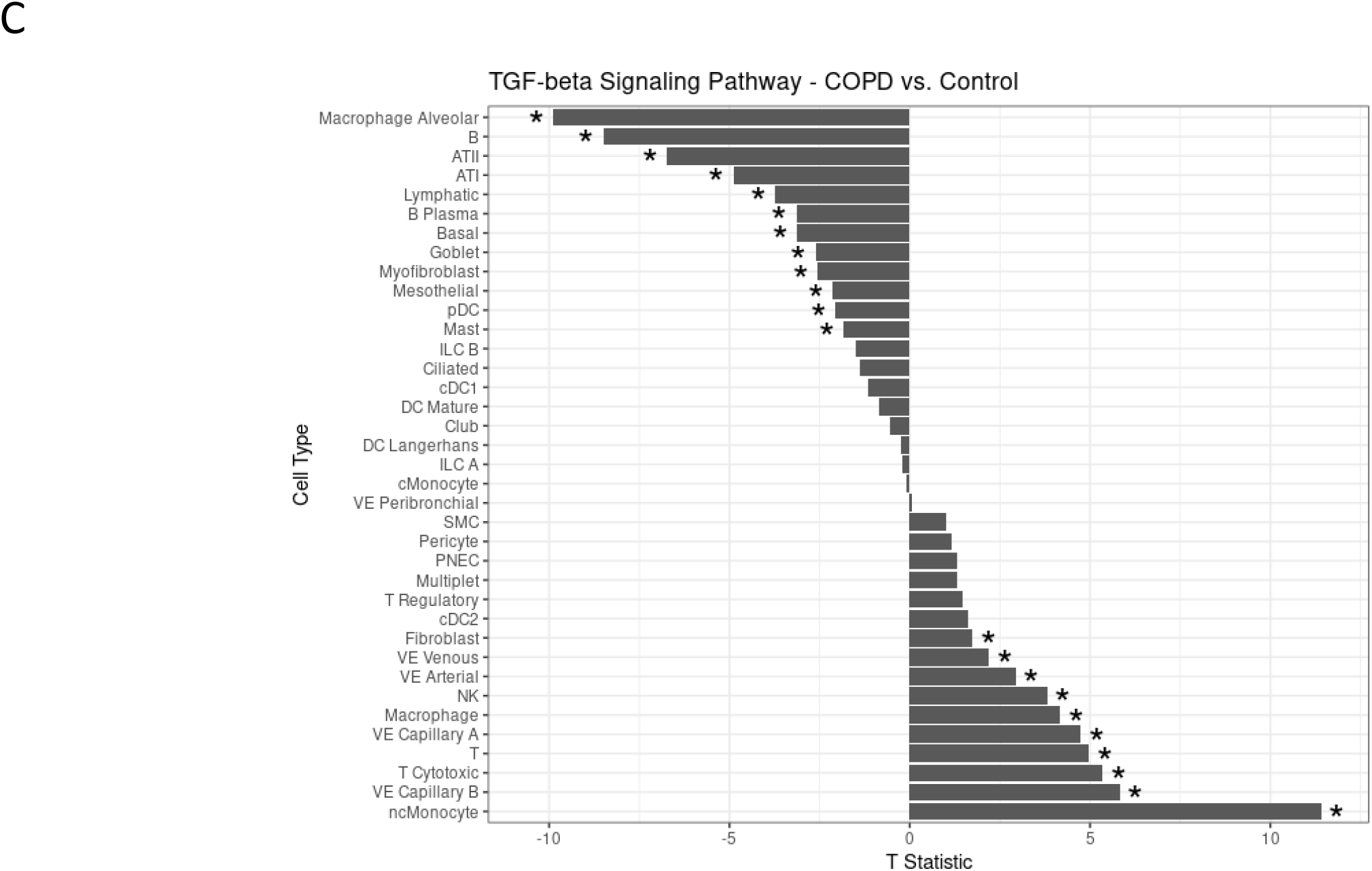
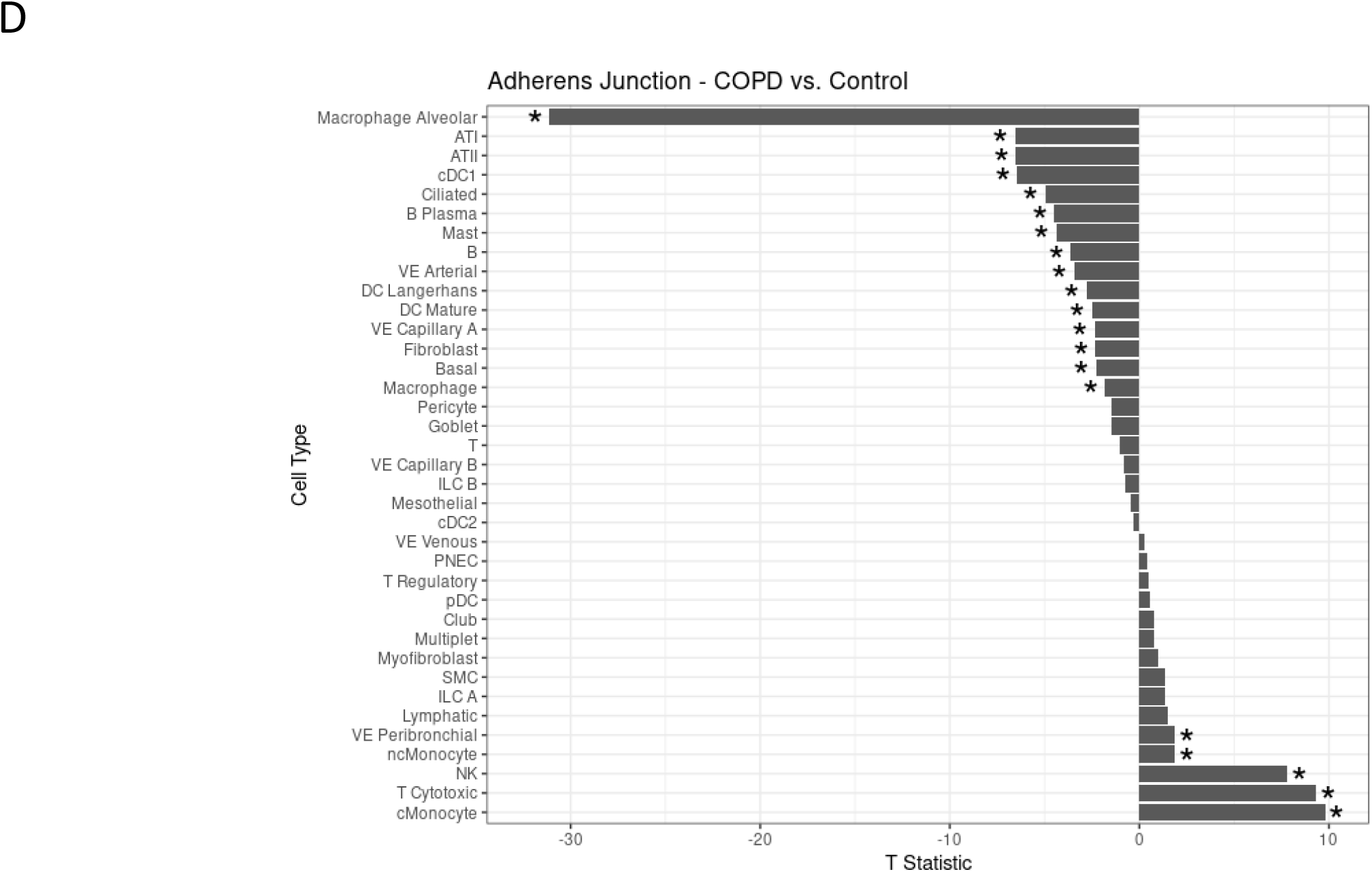
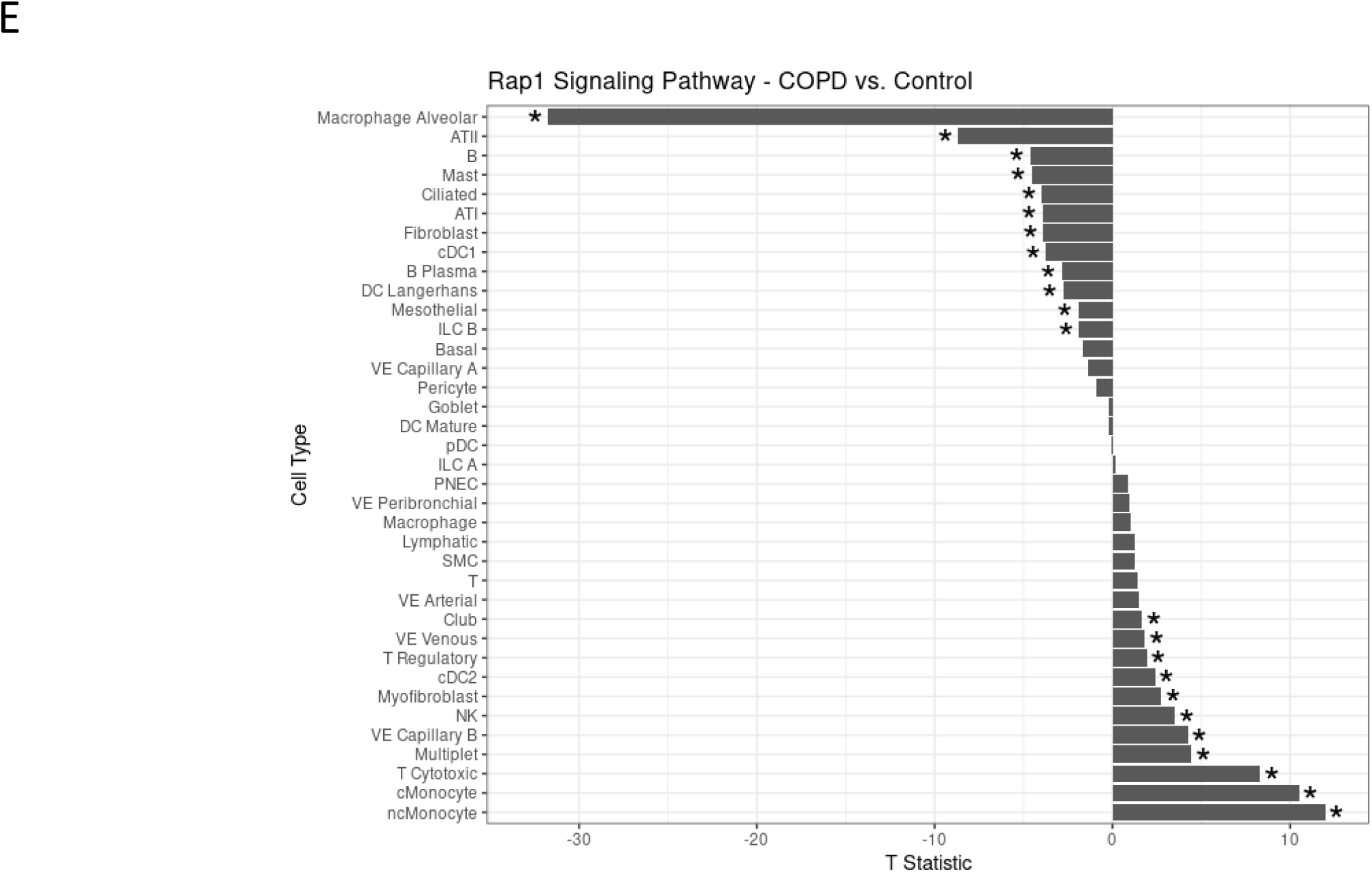
Heatmaps of (**A**) pathway specificity across lung cell types and (**B**) pathway T statistic values of COPD versus control lung tissue samples. T statistic values of COPD versus control lung tissue samples for (**C**) TGF-β signaling pathway, (**D**) adherens junction, and (**E**) Rap1 signaling pathway. Pathway activity scores were generated for each cell in the single-cell dataset. The odds ratio was computed for the cells showing the pathway activity versus not in a given cell type and for the remainder of the data. The P-values were calculated using Fisher’s Exact Test. Additionally, the Welch Two Sample t-test was used to compare the pathway activity scores between controls and end-stage COPD for each cell type separately. A negative T statistic (red) indicates that the pathway has higher activity in COPD compared to controls. A positive T statistic (blue) indicates that the pathway has lower activity in COPD compared to controls. A star indicates that there was a significant difference in activity in COPD versus controls.

To identify cell types showing differential expression of emphysema pathways in subjects with severe COPD, we compared the pathway activity scores between controls and subjects with end-stage COPD for each lung cell type (Figure 4B). Alveolar macrophages had multiple highly dysregulated pathways. The Rap1 signaling, adherens junction, gap junction, and FoxO signaling pathways were more active in alveolar macrophages from subjects with COPD compared to controls. The oxidative phosphorylation and spliceosome pathways were less active in alveolar macrophages from COPD than control subjects. Macrophages, alveolar type (AT) II cells, and B cells clustered together, showing similar changes in pathway activity in COPD compared to controls. Cytotoxic T cells, T cells, NK cells, and non-classical monocytes also clustered together and had decreased activity in COPD samples for nearly every pathway, except for ribosome-associated pathways. When comparing the activity of oxidative phosphorylation in macrophages and monocytes, we observed that macrophages, alveolar macrophages, and classical monocytes (M1 macrophages) had decreased activity, while non-classical monocytes (M2 macrophages) had increased activity.

We then specifically compared the pathway activity scores for the TGF-β signaling pathway, adherens junction, and Rap1 signaling pathway between controls and end-stage COPD for each lung cell type (Figure 4C-E). For all three pathways, there was a disruption in multiple cell types, and cell types were split between having increased or decreased activity. Alveolar macrophages, ATI, ATII, and B cells were among the cell types with the greatest increase in activity in the end-stage COPD compared to controls for all three pathways. Non-classical monocytes and cytotoxic T cells were among the cell types with the greatest decrease in activity in end-stage COPD versus controls. TGF-β signaling was more active in T cells and goblet cells from end-stage COPD samples, while Rap1 and adherens junction signaling were not significantly altered. A list of abbreviations of the cell types can be found in Table E7 in the supplements.

### Comparison of lung and blood transcriptomics

To compare the transcriptomic signature of emphysema in the lung to that in the blood, we obtained data from previously published DGE, DIU, and DEU analyses in whole blood data from COPDGene. There were 247 shared genes among the 1,055 DEGs and 4,913 DEGs from the lung and blood, respectively (Figure 5A). Additionally, 25 DUIs and 2 DUEs were shared between lung and blood samples (Figure 5B-C). GSEA was also run in both lung tissue data from LTRC and whole blood data from COPDGene. The full results from COPDGene can be found in Table E8. When comparing the significant pathways from the blood and lung, there were four shared pathways, two of which were biologically relevant: oxidative phosphorylation and ribosomal RNAs and proteins (Figure 5D and Table E8). Both pathways had negative log-fold changes in both lung and blood, indicating that the majority of genes in the respective gene sets were downregulated with emphysema. The beta coefficients (log-fold changes) of the genes that were significant in either blood or lung were mostly different overall (Figure 5E).

**Figure 5.**
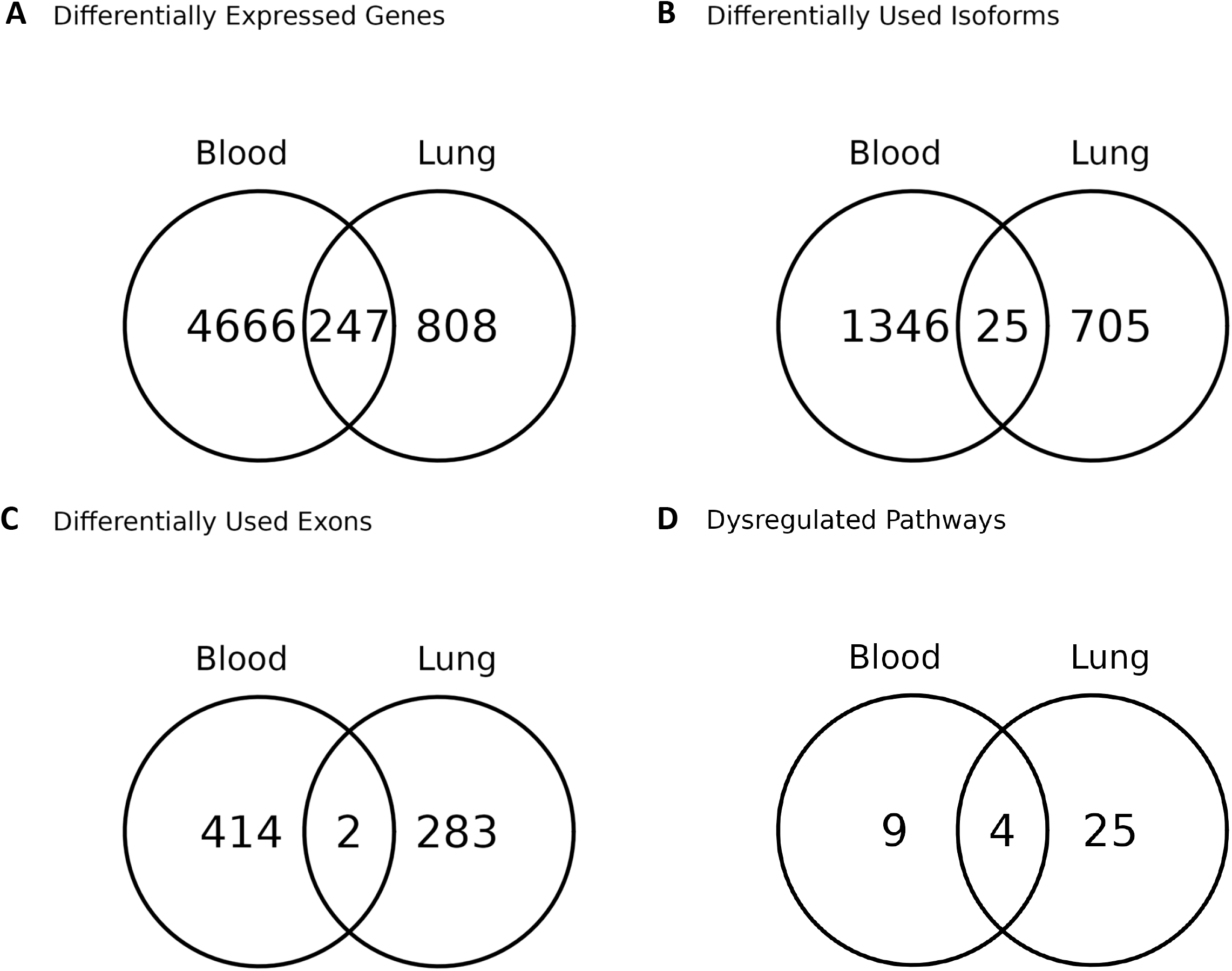
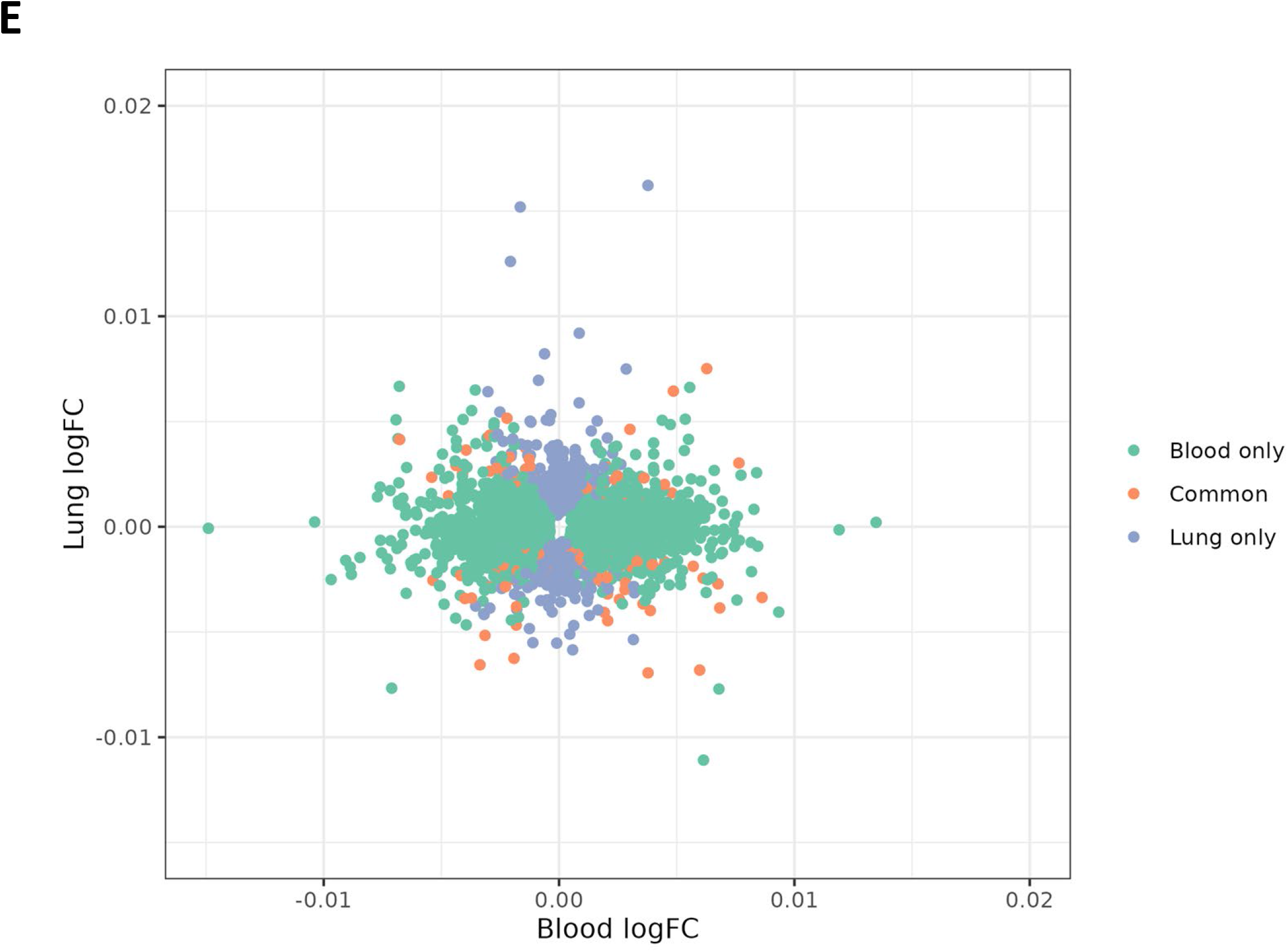
Number of emphysema-associated features shared between blood and lung. (**A**) Differentially expressed genes. (**B**) Differentially used isoforms. (**C**) Differentially used exons. (**D**) Dysregulated pathways. (**E**) Log fold change values of differentially expressed genes associated with emphysema in blood and lung tissue. Genes were included if they were significant in blood, lung tissue, or both *(FDR 10%)*. Emphysema was quantified by Hounsfield units at the 15^th^ percentile of chest CT density histogram at full inspiration (Perc15). The lower the Perc15 values are, i.e. the closer to −1.000 HU, the more CT-quantified emphysema is present. Upregulated versus downregulated pathways are provided with respect to their relationships with Perc15 density which is opposite to emphysema. Negative log fold change values represent upregulated genes and positive log fold change values represent downregulated genes. When comparing the significant pathways from the blood and lung, there were four shared pathways, two of which were biologically relevant: oxidative phosphorylation and ribosomal RNAs and proteins.

## DISCUSSION

In the present study, we identified dysregulated pathways associated with emphysema, which included oxidative phosphorylation, ribosomal function, and pluripotency TGF-β and FoxO signaling pathways. We also found significant associations with pathways involved in epithelial barrier function in multiple cell types. We additionally discovered cell-type specific enrichments, such as decreased and increased oxidative phosphorylation in the pro-inflammatory M1 and anti-inflammatory M2 macrophages, respectively. While there are some shared transcriptomic signals of emphysema in blood and lung tissue, these tissue-specific expression profiles are mostly different.

While prior reports have evaluated the differential gene expression characteristics of emphysema, the knowledge of the alternative splicing mechanisms underlying this disease is limited. Studies have shown that the *SERPINA1* gene, which encodes for the alpha-1 antitrypsin protein involved in the pathophysiology of emphysema in individuals with alpha-1 antitrypsin deficiency, has significant splicing signatures in this disease (53-55). Additionally, the p53/hypoxia-related genes *NUMB* and *PGFA* were significantly associated with alternative splicing signatures in both emphysema and IPF (56). In this transcriptome-wide study of alternative splicing, we identified hundreds of isoforms significantly associated with emphysema. Only 7% of the identified isoforms corresponded to significant genes from the gene-level analysis, indicating the DIU analysis did uncover novel transcriptional events and emphysema-related pathways relative to the standard gene differential expression analysis.

NF-κB is a transcription factor that promotes innate immune and T cell differentiation, suppresses apoptosis, and enhances pro-inflammatory genes (57). Higher amounts of the NF-B p65 subunit protein were found in sputum samples and bronchial biopsies of COPD patients compared to controls (58, 59). We add to these findings by revealing significant associations between emphysema and three NF-κB signaling genes (*PRKCZ, MYBB1A, TRAPPC9)* (60-62). Exposure to cigarette smoke was shown to increase the activity of pathways related to protein kinase C in rat bronchial tissues and mouse alveolar epithelial cells (63, 64). We are the first to demonstrate a relationship between emphysema in human lungs and the *PRKCZ* gene, which generates protein kinase C zeta (65). Reduced MYBBP1A protein levels were previously associated with head and neck squamous cell carcinoma, but no study to date has implicated *MYBB1A* in emphysema (60, 61). We have previously shown an association of *TRAPPC9* with apico-basal emphysema distribution (66), and here we provide more evidence for this association.

Our analysis identified particularly strong and widespread changes in emphysema-associated pathways in alveolar macrophages. Macrophages exposed to oxidants develop an imbalance in the protease/anti-protease activity, particularly in subjects with emphysema who have alpha-1 antitrypsin deficiency (67-69). However, we are the first to show that anti-inflammatory (M2) macrophages (non-classical monocytes) have increased oxidative phosphorylation activity, while pro-inflammatory (M1) macrophages (classical monocytes) have lower activity. M1 macrophages cause tissue damage and block cell division, while M2 macrophages have the opposite effect (70). Cornwell et al. have demonstrated elevated M2 macrophages in COPD (71) but not specifically in emphysema. Together, these findings can be reflective of a process whereby the pro-inflammatory M1 macrophages shift energy metabolism from oxidative phosphorylation to lactic acidosis, and the M2 macrophages predominate in oxygen-dependent metabolism in COPD and emphysema. One study which used single-cell RNA-seq data from explanted human lung tissue showed an elevated expression of the anti-inflammatory and antioxidant-related gene *HMOX1* in advanced COPD (72). Another recent study found that in severe emphysema, macrophages may be responsible for the destruction of bronchiolar and alveolar tissue, as well as the invasion of tertiary lymph nodes (73). This highlights the potential multifaceted nature of macrophages in emphysema, which may warrant future research into this cell type as it relates to emphysema pathophysiology.

Our pathway analysis also revealed enrichment for the pluripotency FoxO signaling and TGF-β signaling pathways. The increased pluripotency function likely preserves cellular homeostasis following lung injury in emphysema, implying a possible role for pluripotent stem cell therapy in emphysema patients (74-76). Both FoxO and TGF-β have been linked to cancer pathogenesis, with the former being a positive mediator of cell growth (77) and vascular remodeling (41) and the latter having both tumor suppressing and promoting functions (78). Studies have shown decreased levels of the FoxO protein in COPD (41, 79-81). Di Stefano et al. found that, compared to controls, COPD peripheral airway samples have decreased TGF-β1^+^ and TGF-β3^+^ bronchial epithelial cells (82). We build on these findings by showing through our lung tissue pathway analysis that TGF-β signaling is dysregulated in emphysema. This finding supports our earlier emphysema integrative genomics and functional investigations, in which we identified functional genetic variations in the *TGFB2* and *ACVR1B* loci in lung fibroblasts and airway epithelial cells (83). The *TGFB2* gene encodes a protein isoform of TGF-β. The *ACVR1B* gene is a transducer of activin-like ligands that are growth and differentiation factors belonging to the TGF-β superfamily of signaling proteins. In another project, we used CRISPR gene editing to discover that the genomic region spanning rs1690789, one of the genetic variants identified by the genome-wide association analysis (GWAS) of emphysema, contains an active enhancer element that increases the expression of TGFB2 in human lung fibroblasts, providing additional evidence of the link between emphysema and TGF-β (84).

With regards to the cell barrier function, damage to the alveolar respiratory epithelium has been shown to be linked to emphysema and COPD (85, 86). We showed that a wide array of cell types is significantly associated with TGF-β signaling, adherens junction, and Rap1 signaling pathways, all related to epithelial barrier homeostasis, in emphysema and end-stage COPD. For example, alveolar macrophages, B cells, plasma cells, pulmonary ATI and ATII cells are significantly downregulated, while natural killer and T lymphocytes are significantly upregulated for all three pathways.

The current study has a number of strengths. It is the largest study to date to investigate the relationship of emphysema to gene expression and alternative splicing. Pathway enrichment analysis revealed novel biological processes and confirmed the results of previously published COPD and emphysema studies. In addition, by integrating our bulk RNA-seq analysis with another single-cell dataset from subjects with severe COPD, we were able to identify specific lung cell types showing COPD-associated dysregulation of the pathways identified in the bulk tissue analysis.

There are also limitations to this study. One limitation is that we did not include a sizable number of LTRC participants in our analyses. Most samples were excluded due to missing CT emphysema values. An additional limitation is that the single-cell dataset we queried was made up of patients with end-stage COPD rather than emphysema specifically. However, emphysema is typically present in end-stage COPD patients (87). Future research should examine enriched pathways using bulk and single-cell RNA-seq data from the same cohorts to shed further light on the various cell types implicated in emphysema pathobiology. Lastly, the large sample size for our primary analysis reduces the risk of false positive associations, but further validation of these results in comparable cohorts will provide greater confidence in these associations.

## CONCLUSION

CT-quantified emphysema exhibits distinctive alternative splicing and transcriptomic patterns, as well as associations to biological processes connected to oxidative phosphorylation and pluripotency pathways. In addition to other cell-type-specific markers, the M1/M2 macrophage ratio was found to be decreased in relation to oxidative phosphorylation, and a number of innate and adaptive immune cell types are enriched for epithelial barrier function. These novel molecular and cellular findings in emphysema may help in the development of effective screening methods and therapeutic strategies.

## Data Availability

All data produced in the present study are available upon reasonable request to the authors.

## COPDGene Investigators - Core Units

### Administrative Center

James D. Crapo, MD (PI); Edwin K. Silverman, MD, PhD (PI); Barry J. Make, MD; Elizabeth A. Regan, MD, PhD

### Genetic Analysis Center

Terri H. Beaty, PhD; Peter J. Castaldi, MD, MSc; Michael H. Cho, MD, MPH; Dawn L. DeMeo, MD, MPH; Adel Boueiz, MD, MMSc; Marilyn G. Foreman, MD, MS; Auyon Ghosh, MD; Lystra P. Hayden, MD, MMSc; Craig P. Hersh, MD, MPH; Jacqueline Hetmanski, MS; Brian D. Hobbs, MD, MMSc; John E. Hokanson, MPH, PhD; Wonji Kim, PhD; Nan Laird, PhD; Christoph Lange, PhD; Sharon M. Lutz, PhD; Merry-Lynn McDonald, PhD; Dmitry Prokopenko, PhD; Matthew Moll, MD, MPH; Jarrett Morrow, PhD; Dandi Qiao, PhD; Elizabeth A. Regan, MD, PhD; Aabida Saferali, PhD; Phuwanat Sakornsakolpat, MD; Edwin K. Silverman, MD, PhD; Emily S. Wan, MD; Jeong Yun, MD, MPH

### Imaging Center

Juan Pablo Centeno; Jean-Paul Charbonnier, PhD; Harvey O. Coxson, PhD; Craig J. Galban, PhD; MeiLan K. Han, MD, MS; Eric A. Hoffman, Stephen Humphries, PhD; Francine L. Jacobson, MD, MPH; Philip F. Judy, PhD; Ella A. Kazerooni, MD; Alex Kluiber; David A. Lynch, MB; Pietro Nardelli, PhD; John D. Newell, Jr., MD; Aleena Notary; Andrea Oh, MD; Elizabeth A. Regan, MD, PhD; James C. Ross, PhD; Raul San Jose Estepar, PhD; Joyce Schroeder, MD; Jered Sieren; Berend C. Stoel, PhD; Juerg Tschirren, PhD; Edwin Van Beek, MD, PhD; Bram van Ginneken, PhD; Eva van Rikxoort, PhD; Gonzalo Vegas SanchezFerrero, PhD; Lucas Veitel; George R. Washko, MD; Carla G. Wilson, MS

### PFT QA Center, Salt Lake City, UT

Robert Jensen, PhD

### Data Coordinating Center and Biostatistics, National Jewish Health, Denver, CO

Douglas Everett, PhD; Jim Crooks, PhD; Katherine Pratte, PhD; Matt Strand, PhD; Carla G. Wilson, MS

### Epidemiology Core, University of Colorado Anschutz Medical Campus, Aurora, CO

John E. Hokanson, MPH, PhD; Erin Austin, PhD; Gregory Kinney, MPH, PhD; Sharon M. Lutz, PhD; Kendra A. Young, PhDVersion Date: March 26, 2021

### Mortality Adjudication Core

Surya P. Bhatt, MD; Jessica Bon, MD; Alejandro A. Diaz, MD, MPH; MeiLan K. Han, MD, MS; Barry Make, MD; Susan Murray, ScD; Elizabeth Regan, MD; Xavier Soler, MD; Carla G. Wilson, MS

### Biomarker Core

Russell P. Bowler, MD, PhD; Katerina Kechris, PhD; Farnoush BanaeiKashani, PhD

## COPDGene Investigators - Clinical Centers

### Ann Arbor VA

Jeffrey L. Curtis, MD; Perry G. Pernicano, MD

### Baylor College of Medicine, Houston, TX

Nicola Hanania, MD, MS; Mustafa Atik, MD; Aladin Boriek, PhD; Kalpatha Guntupalli, MD; Elizabeth Guy, MD; Amit Parulekar, MD

### Brigham and Women’s Hospital, Boston, MA

Dawn L. DeMeo, MD, MPH; Craig Hersh, MD, MPH; Francine L. Jacobson, MD, MPH; George Washko, MD

### Columbia University, New York, NY

R. Graham Barr, MD, DrPH; John Austin, MD; Belinda D’Souza, MD; Byron Thomashow, MD

### Duke University Medical Center, Durham, NC

Neil MacIntyre, Jr., MD; H. Page McAdams, MD; Lacey Washington, MD

### HealthPartners Research Institute, Minneapolis, MN

Charlene McEvoy, MD, MPH; Joseph Tashjian, MD

### Johns Hopkins University, Baltimore, MD

Robert Wise, MD; Robert Brown, MD; Nadia N. Hansel, MD, MPH; Karen Horton, MD; Allison Lambert, MD, MHS; Nirupama Putcha, MD, MHS

### Lundquist Institute for Biomedical Innovation at Harbor UCLA Medical Center, Torrance, CA

Richard Casaburi, PhD, MD; Alessandra Adami, PhD; Matthew Budoff, MD; Hans Fischer, MD; Janos Porszasz, MD, PhD; Harry Rossiter, PhD; William Stringer, MD

### Michael E. DeBakey VAMC, Houston, TX

Amir Sharafkhaneh, MD, PhD; Charlie Lan, DO

### Minneapolis VA

Christine Wendt, MD; Brian Bell, MD; Ken M. Kunisaki, MD, MS

### Morehouse School of Medicine, Atlanta, GA

Eric L. Flenaugh, MD; Hirut Gebrekristos, PhD; Mario Ponce, MD; Silanath Terpenning, MD; Gloria Westney, MD, MS

### National Jewish Health, Denver, CO

Russell Bowler, MD, PhD; David A. Lynch, MB

### Reliant Medical Group, Worcester, MA

Richard Rosiello, MD; David Pace, MD

### Temple University, Philadelphia, PA

Gerard Criner, MD; David Ciccolella, MD; Francis Cordova, MD; Chandra Dass, MD; Gilbert D’Alonzo, DO; Parag Desai, MD; Michael Jacobs, PharmD; Steven Kelsen, MD, PhD; Victor Kim, MD; A. James Mamary, MD; Nathaniel Marchetti, DO; Aditi Satti, MD; Kartik Shenoy, MD; Robert M. Steiner, MD; Alex Swift, MD; Irene Swift, MD; Maria Elena Vega-Sanchez, MD

### University of Alabama, Birmingham, AL

Mark Dransfield, MD; William Bailey, MD; Surya P. Bhatt, MD; Anand Iyer, MD; Hrudaya Nath, MD; J. Michael Wells, MD

### University of California, San Diego, CA

Douglas Conrad, MD; Xavier Soler, MD, PhD; Andrew Yen, MD

### University of Iowa, Iowa City, IA

Alejandro P. Comellas, MD; Karin F. Hoth, PhD; John Newell, Jr., MD; Brad Thompson, MD

### University of Michigan, Ann Arbor, MI

MeiLan K. Han, MD MS; Ella Kazerooni, MD MS; Wassim Labaki, MD MS; Craig Galban, PhD; Dharshan Vummidi, MD

### University of Minnesota, Minneapolis, MN

Joanne Billings, MD; Abbie Begnaud, MD; Tadashi Allen, MD

### University of Pittsburgh, Pittsburgh, PA

Frank Sciurba, MD; Jessica Bon, MD; Divay Chandra, MD, MSc; Joel Weissfeld, MD, MPH

### University of Texas Health, San Antonio, San Antonio, TX

Antonio Anzueto, MD; Sandra Adams, MD; Diego Maselli-Caceres, MD; Mario E. Ruiz, MD; Harjinder Singh

