## Supplement for "Lung transcriptomics of radiologic emphysema reveal barrier function impairment and macrophage M1-M2 imbalance"

Robin Lu^1^+, Andrew Gregory^1^+, Rahul Suryadevara^1^, Zhonghui Xu^1^, Dhawal Jain^1,2^, Brian D. Hobbs^1,3^, Noah Lichtblau^1^, Robert Chase^1^, Edwin K. Silverman^1,3^, Craig P. Hersh^1,3^, Peter J. Castaldi^1,4^*, Adel Boueiz^1,3^*, for the COPDGene investigators.

+ Equal contribution

* Equal contribution

^1^Channing Division of Network Medicine, Brigham and Women’s Hospital, Harvard Medical School, Boston, MA; ^2^Division of Pulmonary Drug Discovery Laboratory, Bayer US LLC. Pharmaceuticals, Research and Development, Boston, MA; ^3^Division of Pulmonary and Critical Care Medicine, Brigham and Women’s Hospital, Harvard Medical School, Boston, MA; ^4^Division of General Medicine and Primary Care, Brigham and Women’s Hospital, Harvard Medical School, Boston, MA.

**Supplemental methods**

***RNA extraction, quality control, and alignment***

RNA quantification for LTRC was performed using the Quant-iT RNA assay (Invitrogen) and RNA integrity analysis using a fragment analyzer (Advanced Analytical). Samples with low RNA total amount, concentration, or integrity were rejected. The TruSeq Stranded mRNA kit (Illumina) was used to perform poly-A selection and cDNA synthesis. Final RNA-seq libraries were quantified using the Quant-it dsDNA High Sensitivity assay, and base calls were generated in real-time on the NovaSeq6000 instrument (RTA 3.1.5). For COPDGene, total blood RNA was extracted from subjects at Visit 2 in PAXgene TM Blood RNA tubes from the Qiagen PreAnalytiX PAXgene Blood miRNA Kit (Qiagen, Valencia, CA).

Paired-end reads were generated from Illumina sequencers. Alignment to GRCh38 was performed using STAR(1). Gene Transfer Format (GTF) annotation was downloaded from GENCODE release 37. Exons from the GTF were broken into disjoint parts (exonic parts) sharing a common set of transcripts (2). The featureCounts function in the Rsubread package was used to generate sequencing read counts for exonic parts (v2.4.3) (3). Gene and isoform expression estimates were obtained using Salmon (v1.3.0) (4) and summarized to gene level using tximeta (v1.8.5) (5). Sex consistency was confirmed by comparing the expression of Xist and non-pseudo-autosomal regions of the Y chromosome. The gene and isoform count data used for this analysis are available in the Gene Expression Omnibus (6, 7).

***Filtering and normalization***

Genomic features with less than one count per million reads in more than 20% of subjects were filtered out prior to applying trimmed mean of M values normalization from edgeR (v3.32.1), which accounts for differences in sequencing depth and library composition (8).

**SUPPLEMENTARY FIGURE LEGENDS**

**Figure E1.** Missing covariates for excluded Lung Tissue Research Consortium (LTRC) samples. Abbreviations: BMI: Body mass index. FEV_1_: Forced expiratory volume in 1 second. Total Perc15 density: Hounsfield units at the 15^th^ percentile of CT density histogram at total lung capacity.

**Figure E2.** Selection of COPDGene study subjects. Abbreviations: AA: African American. Adjusted Perc15 density: Hounsfield units at the 15^th^ percentile of CT density histogram at total lung capacity corrected for the inspiratory depth variations. BMI: Body mass index. CBC: Complete blood count. FEV_1_: Forced expiratory volume in 1 second. IPF: Idiopathic pulmonary fibrosis. NHW: Non-Hispanic white.

**Figure E3.** Distribution of (**A**) total Perc15 density in the Lung Tissue Research Consortium (LTRC) subjects and (**B**) adjusted Perc15 density in the COPDGene subjects. Abbreviations: Total Perc15 density: Hounsfield units at the 15^th^ percentile of CT density histogram at total lung capacity. Adjusted Perc15 density: Hounsfield units at the 15^th^ percentile of CT density histogram at total lung capacity corrected for the inspiratory depth variations. Both total Perc15 and adjusted Perc15 density are given as the Hounsfield units + 1000.

**Figure E4.** KEGG (Kyoto Encyclopedia of Genes and Genomes) pathway maps of the (**A**) oxidative phosphorylation, (**B)** signaling pathways regulating pluripotency of stem cells, (**C)** regulation of actin cytoskeleton, (**D)** JAK-STAT signaling pathway, (**E)** Fc epsilon RI signaling pathway, and (**F)** gap junction reporting the effect size (log fold change) of all genes within each pathway in the 456 lung tissue samples from subjects in the Lung Tissue Research Consortium (LTRC). Emphysema was quantified by Hounsfield units at the 15^th^ percentile of chest CT density histogram at full inspiration (Perc15). The lower the Perc15 values are, i.e. the closer to -1,000 HU, the more CT-quantified emphysema is present. Gene log fold changes were multiplied by -100 so that positive log fold changes represented upregulated genes and negative log fold changes represented downregulated genes. Red represents upregulated genes and blue represents downregulated genes.
