## Supplementary figures and images for "Lung transcriptomics of radiologic emphysema reveal barrier function impairment and macrophage M1-M2 imbalance"

### Supplementary figures

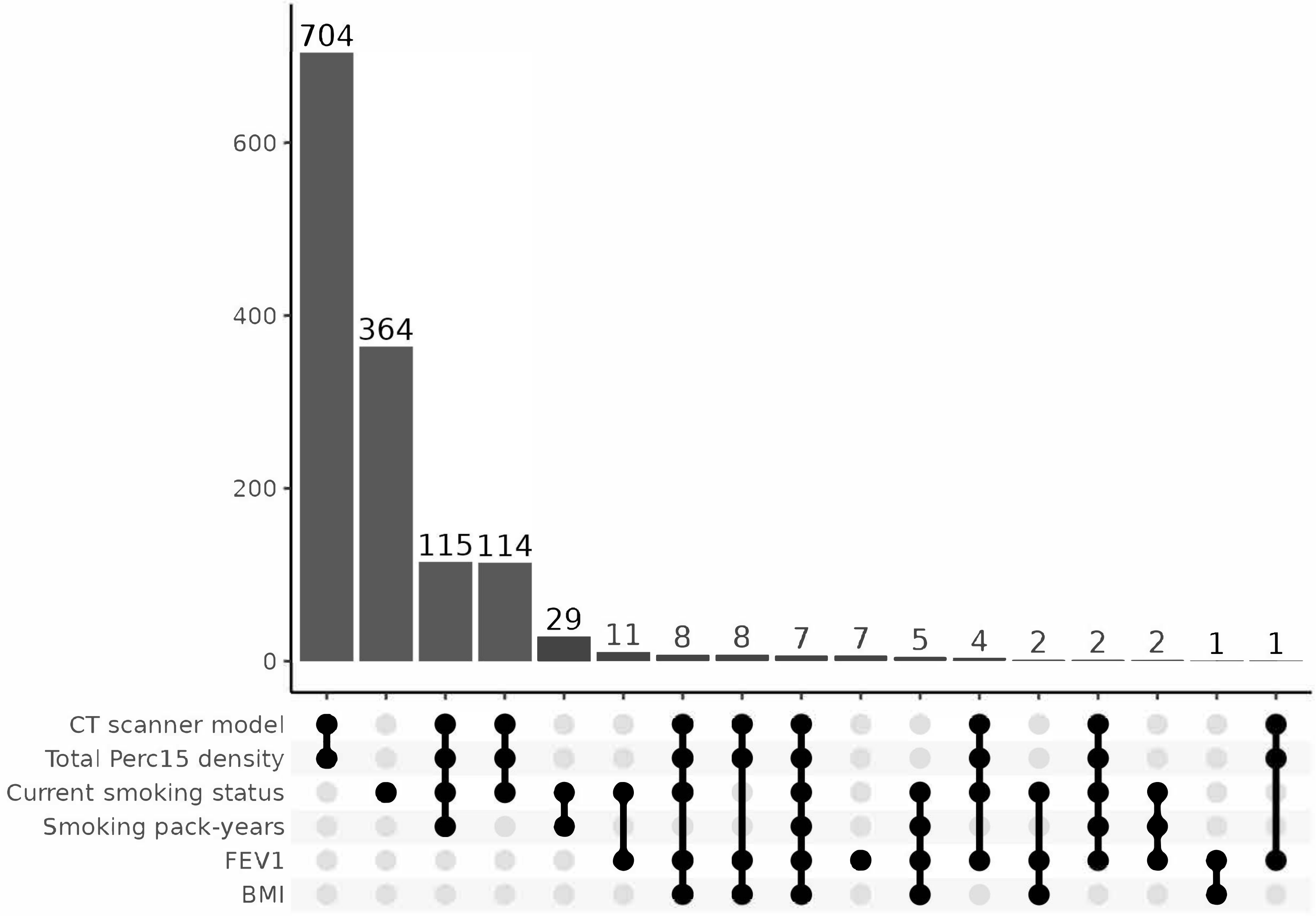

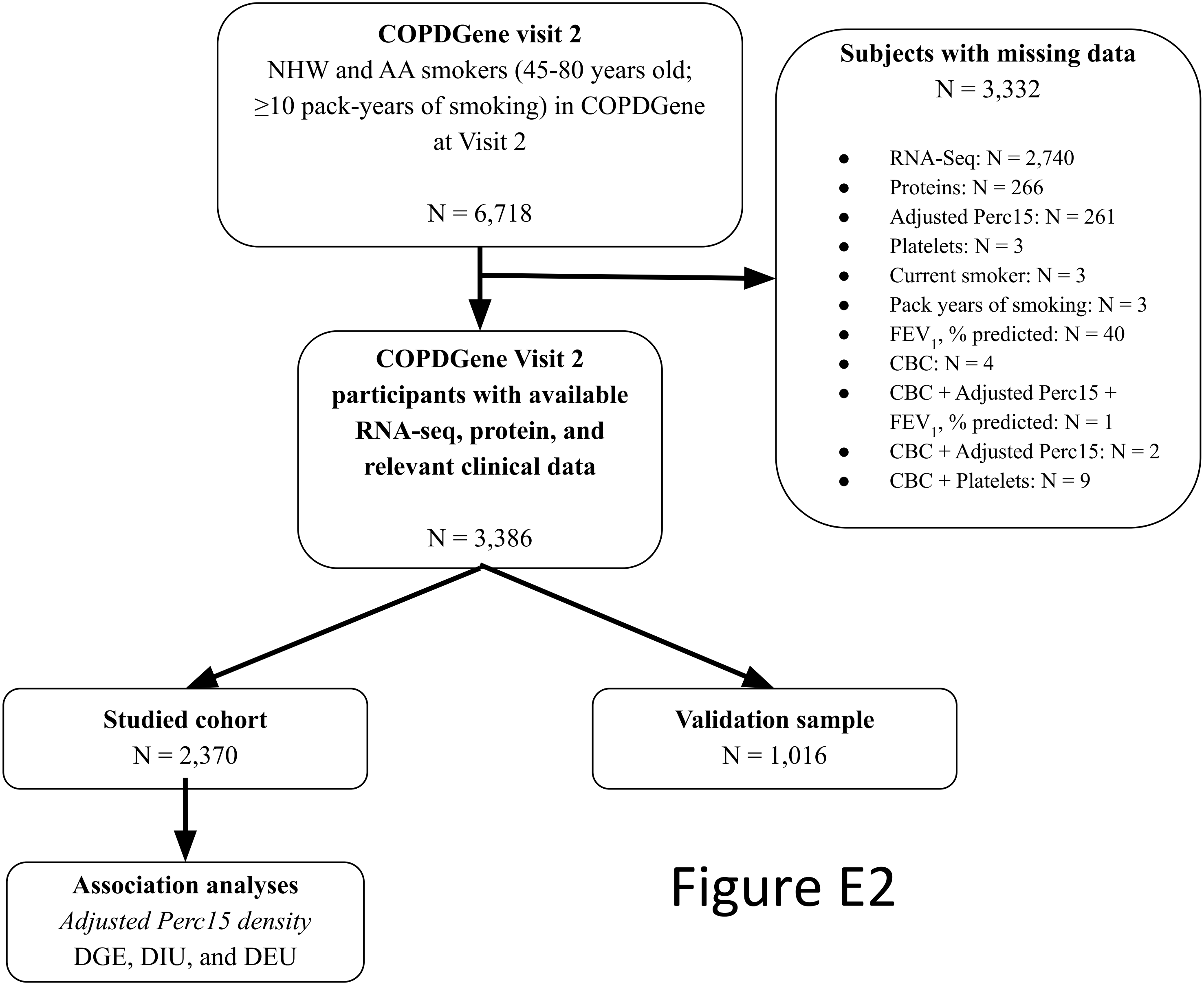

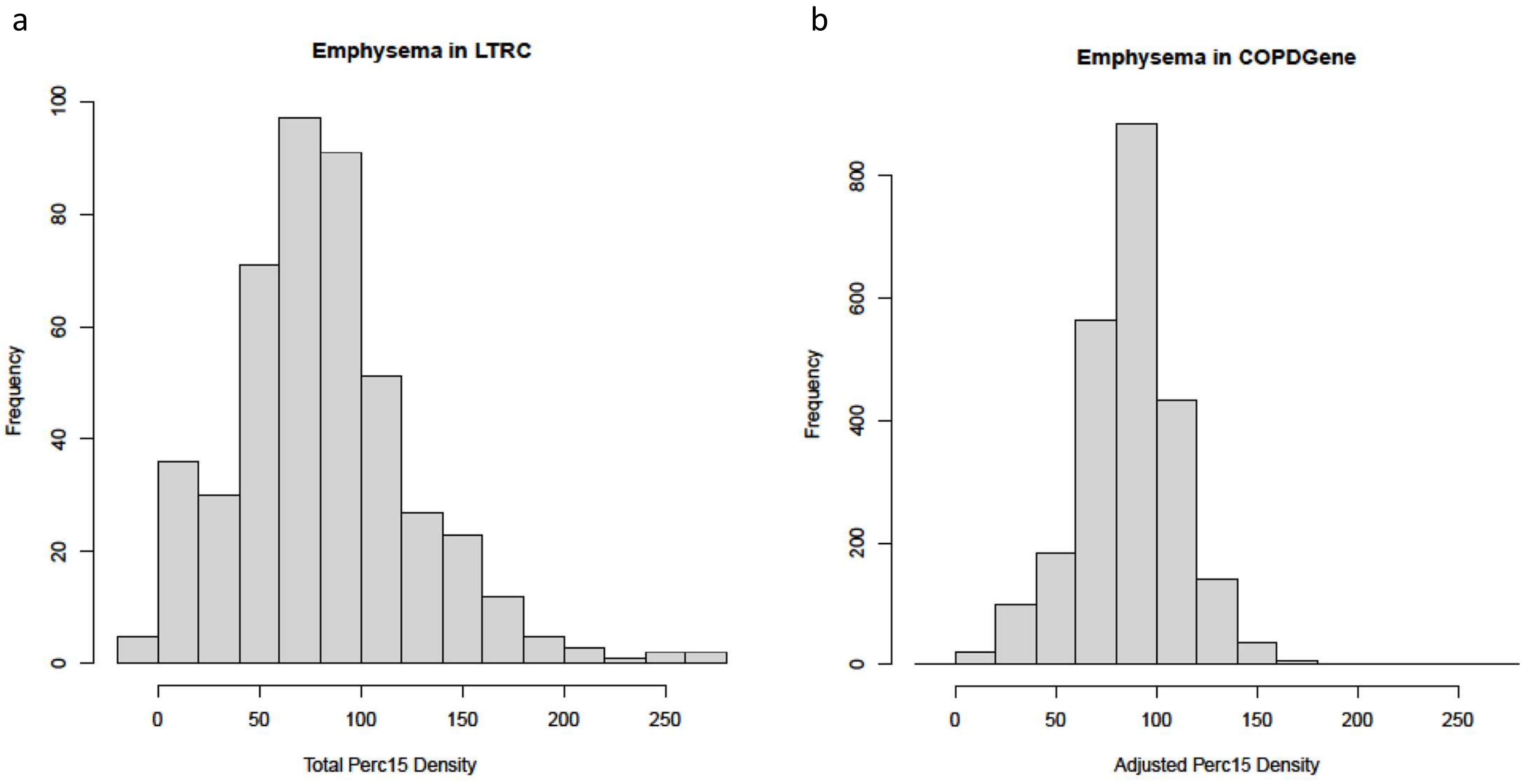

#### Figure E4

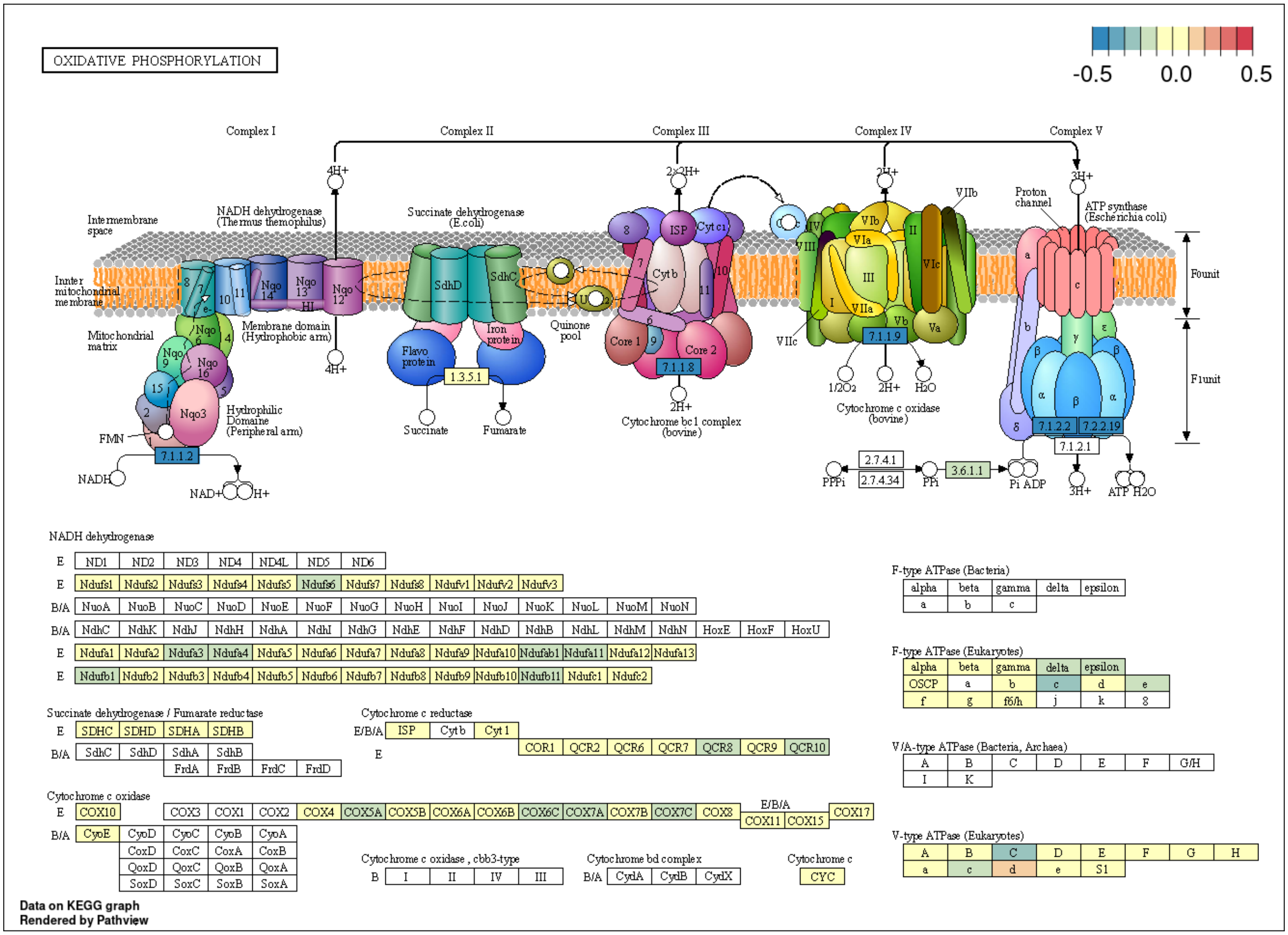

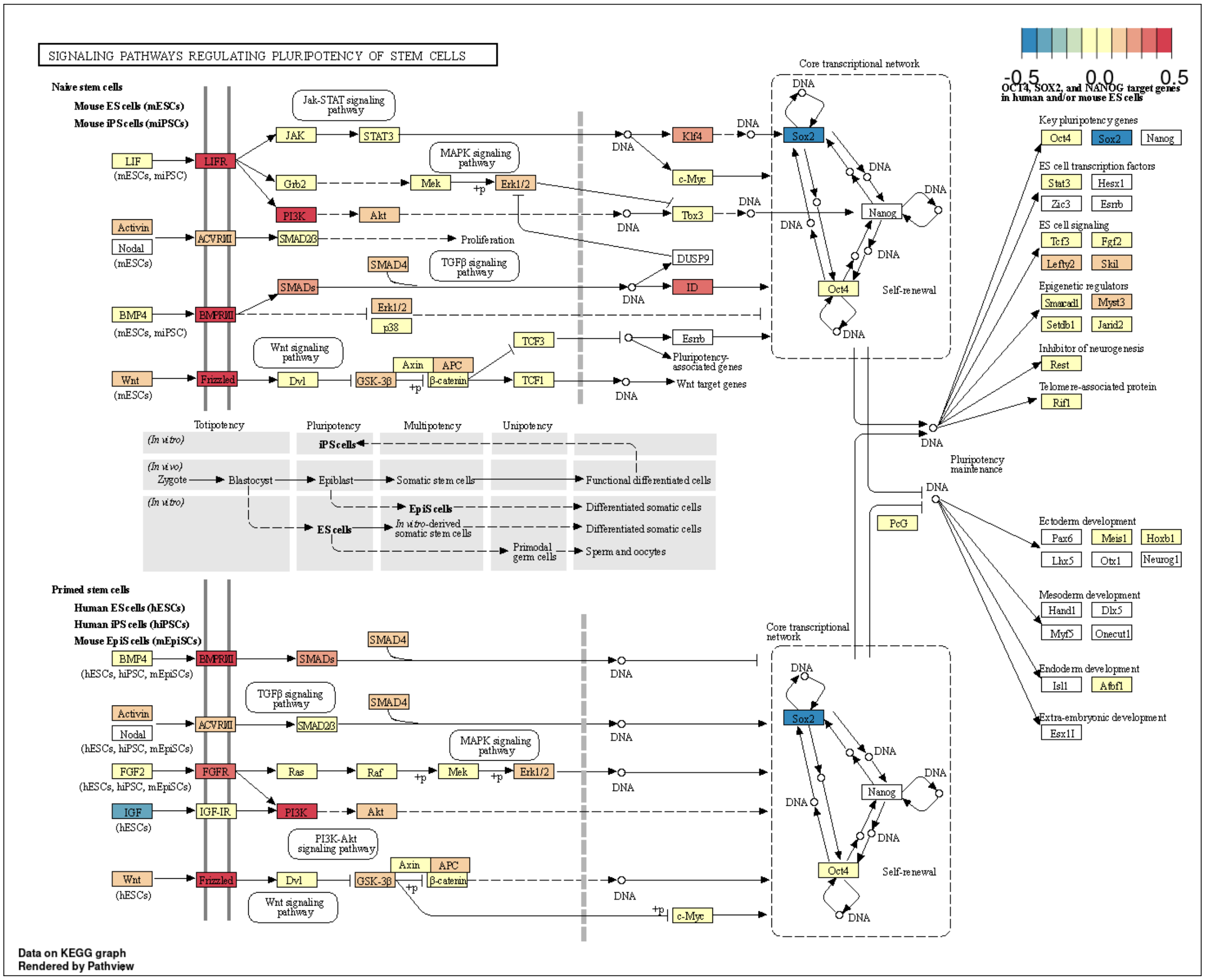

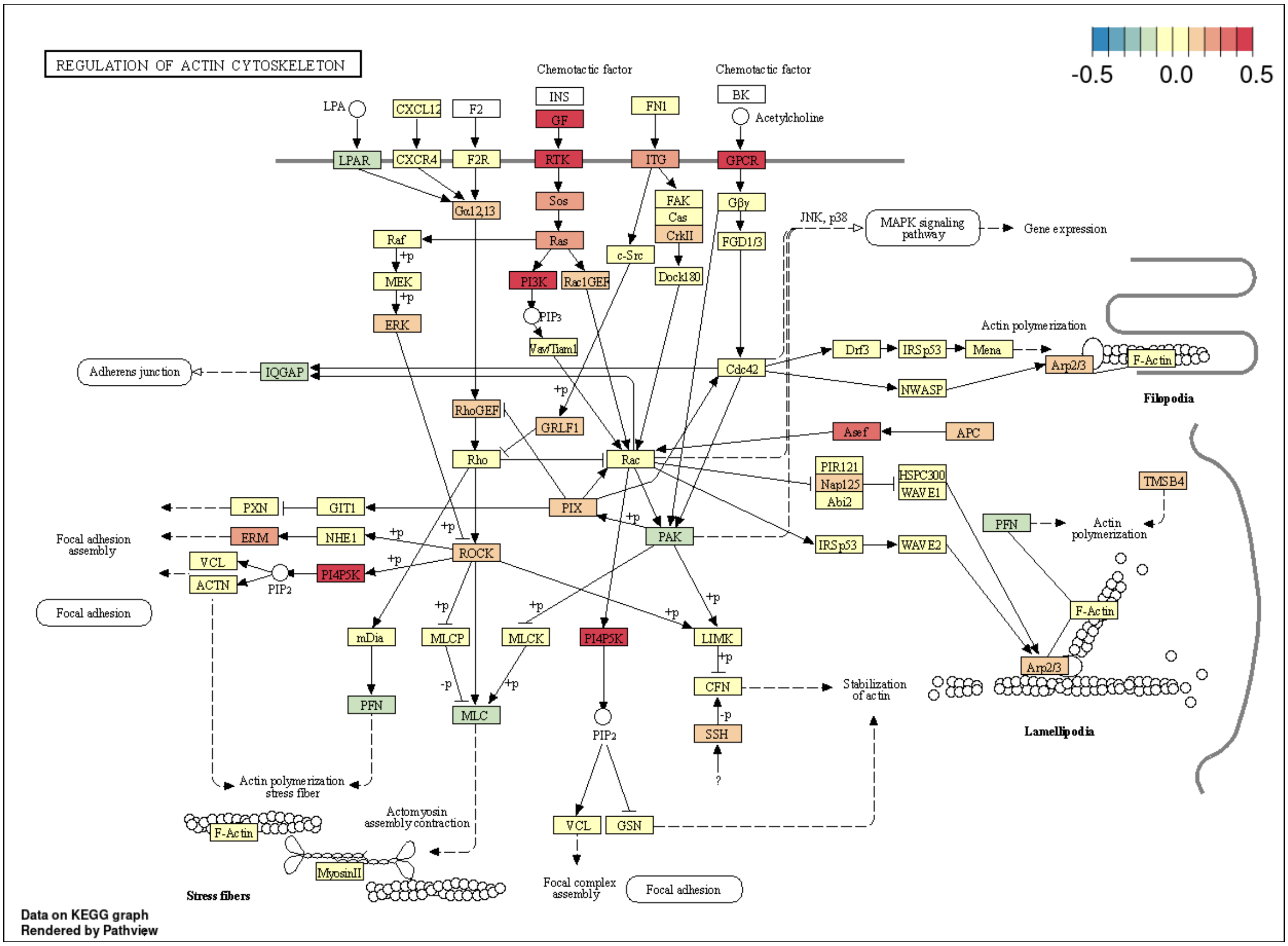

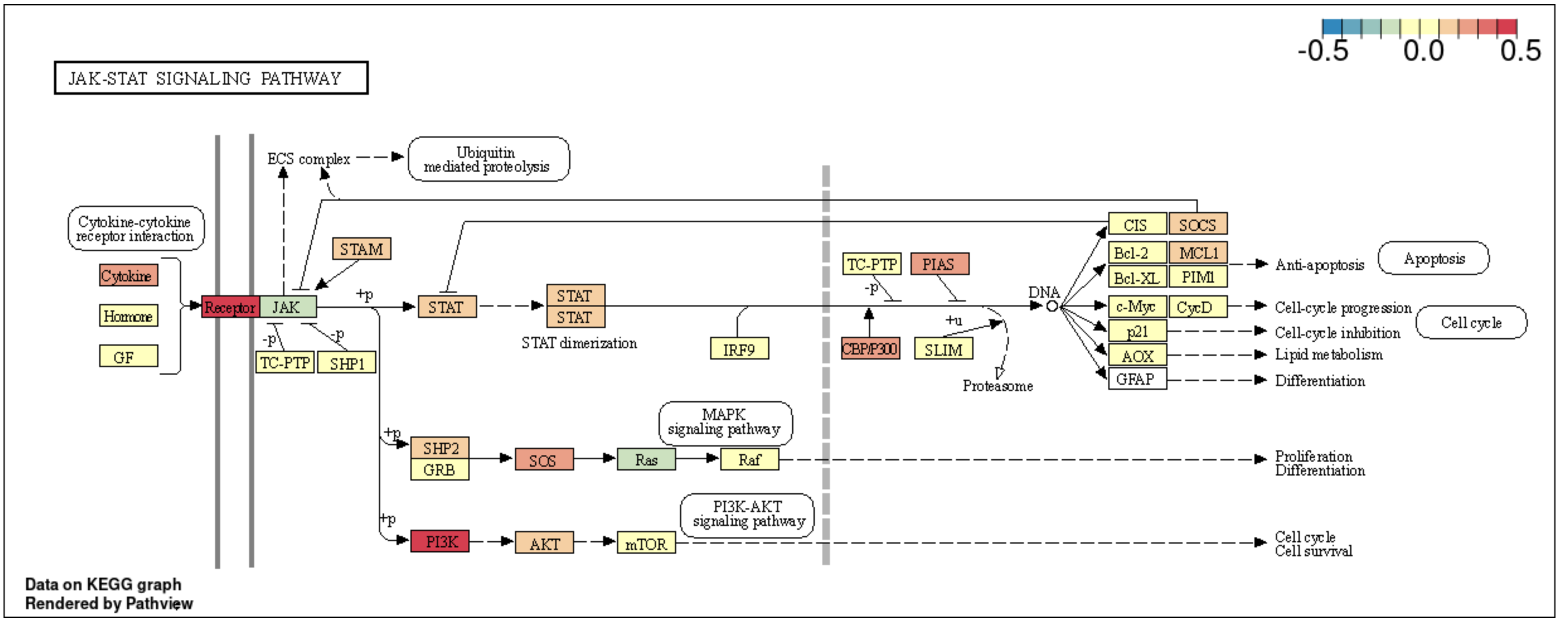

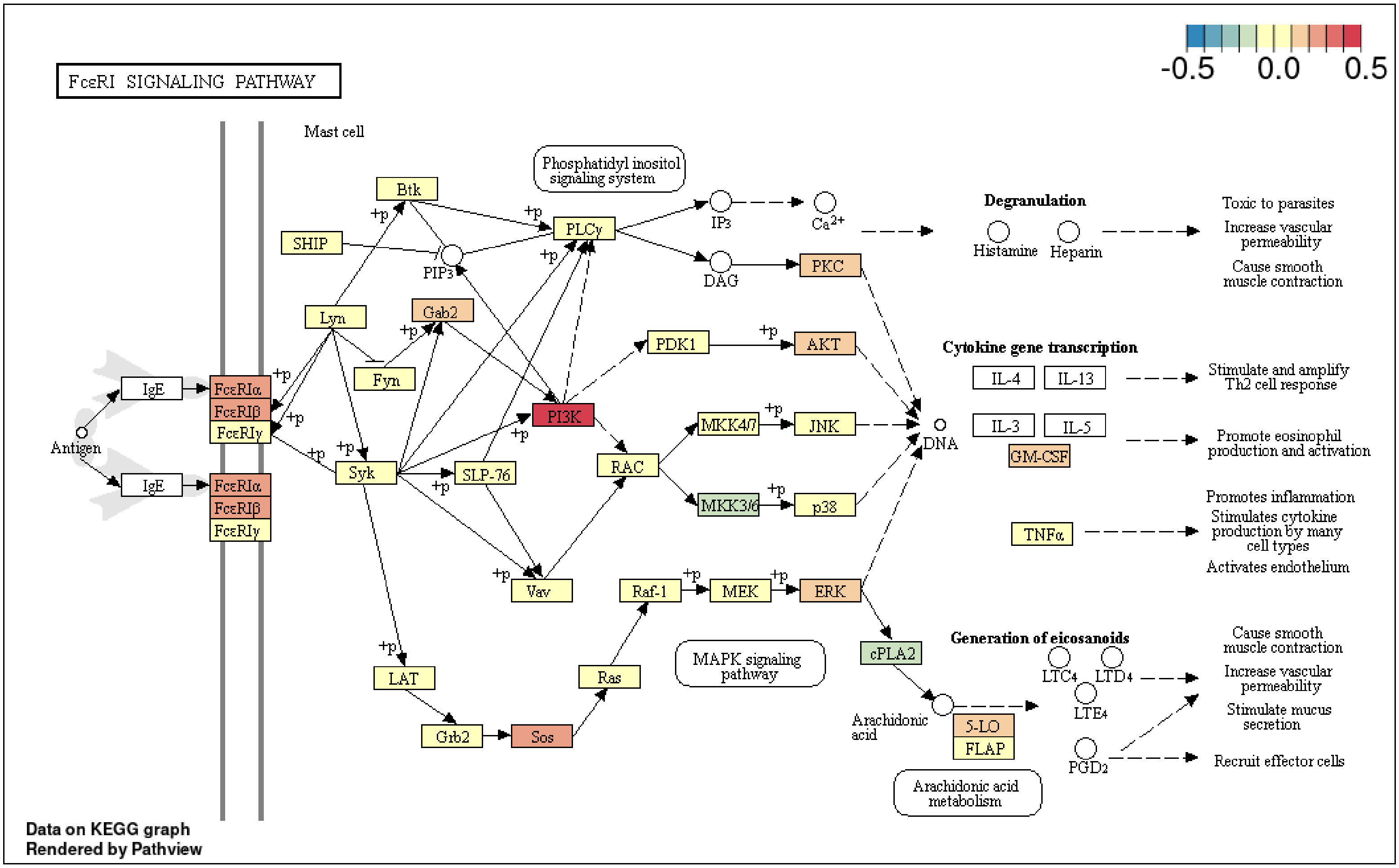

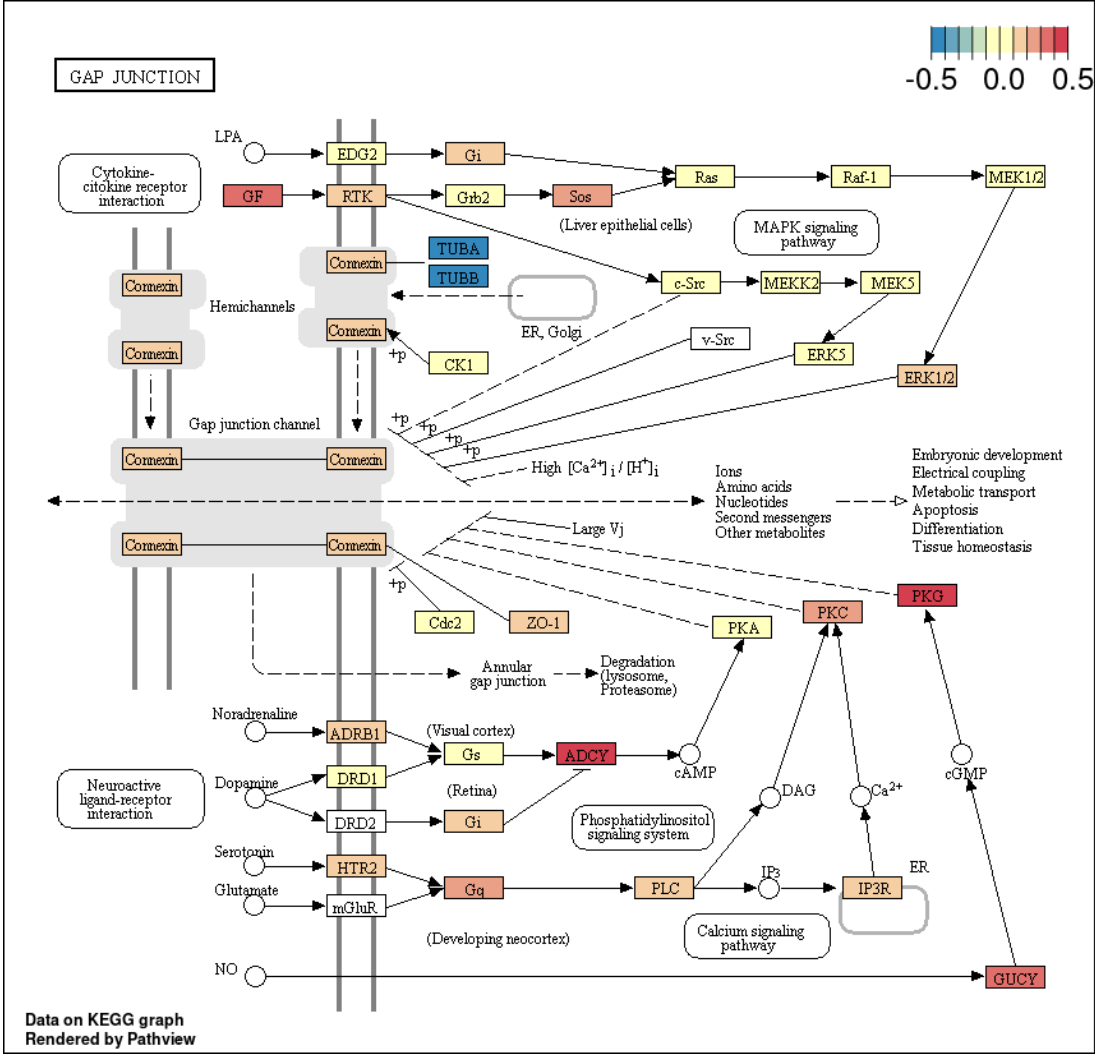
